# A phase I window-of-opportunity study of neoadjuvant avelumab and hypofractionated radiation therapy in radiation-relapsed meningioma

**DOI:** 10.1101/2025.09.26.25336740

**Authors:** Jiayi Huang, Tanner M. Johanns, Jian L. Campian, Michael Chicoine, Gregory Zipfel, Milan Chheda, Omar Butt, Danil Ivanov, Konstantin Chernyshov, Basim Salem, Arina Tkachuk, Alena Frank, Daria Klimova, Dmitry Tabakov, Akdes S. Harmanci, William C. Chen, Akash J. Patel, David R. Raleigh, Albert H. Kim, Chun-Kan Chen

## Abstract

Effective treatments for recurrent, radiation-relapsed meningiomas (RR-meningiomas) following surgery and radiation therapy (RT) are limited. Inhibitors of the programmed-death-1 (PD-1) or programmed-death ligand-1 (PD-L1) pathway have shown modest activity in single-arm phase II studies of RR-meningiomas. This study aimed to evaluate the immunological effects of combining avelumab, a PD-L1 inhibitory antibody, with proton beam therapy (PBT) in RR-meningiomas. Patients with grade 1-3 RR-meningiomas were treated with neoadjuvant avelumab plus hypofractionated PBT, followed by surgery and adjuvant avelumab. Correlative analyses included RNA-sequencing (RNA-seq), whole exome sequencing (WES), multiplex immunofluorescence (MxIF), single-nucleus RNA-seq (snRNA-seq) of pre- and post-treatment tumor tissues, and flow cytometry (FC) of serial blood samples. Nine patients were enrolled: three achieved an immunologic response and prolonged progression-free survival (PFS > 36 months). At a median follow-up of 47.2 months, the median PFS was 19.1 months (95% CI: 15.2-23.0). RNA-seq showed a dynamic change of tumor microenvironment (TME) signatures. MxIF revealed marked infiltration of T cells and CD68^+^CD206^−^ (M1-phenotype) macrophages in the post-treatment tissues of responders, a pattern absent in non-responders, whose pre- and post-treatment tissues predominantly featured CD206^+^ (M2-phenotype) macrophages. These findings were supported by snRNA-seq, which identified FN1-associated immunosuppressive macrophage subtype enriched in non-responders. Additionally, FC revealed elevated peripheral primed T-cell signatures one-month after treatment initiation in responders, suggesting a potential predictive biomarker. Avelumab combined with RT may elicit an immune response in a subset of RR-meningiomas, leading to prolonged remission. Further investigations are warranted to validate these findings and to develop predictive biomarkers in larger prospective studies.

**Summary:** This phase I window-of-opportunity trial demonstrates that neoadjuvant PD-L1 blockade combined with hypofractionated proton therapy can induce robust immunologic responses and prolonged progression-free survival in a subset of patients with radiation-relapsed meningioma, supported by multi-omic profiling of the tumor microenvironment and peripheral blood.

## INTRODUCTION

Meningioma is the most common central nervous system (CNS) tumor, accounting for approximately 42% of all CNS tumors and an estimated 39,000 newly cases annually in the United States (*1*). Although most meningiomas follow a relatively indolent course and can be managed with observation, surgery, or radiation therapy (RT), 10-25% of grade 1 meningiomas and 30-50% of grade 2-3 meningiomas recur despite surgery and RT (*2, 3*). For patients with recurrent radiation-relapsed meningiomas (RR-meningiomas) that progressed after surgery and RT, no effective systemic therapy exists. In a comprehensive review of systemic therapies for RR-meningiomas, the 6-month progression-free survival (PFS-6) remained poor across World Health Organization [WHO] grades. The weighted average PFS-6 of WHO grade I was 29% (95% CI: 20%– 38%), and the weighted average PFS-6 WHO grade II/III was 26% (95% CI: 19%–33%) (*4*). In the absence of effective systemic therapies, repeat surgery or reirradiation is often pursued (*5, 6*), including the use of proton beam therapy (PBT) (*7*). However, these salvage strategies offer limited durable disease control, underscoring the need for novel therapeutic approaches.

Programmed-death ligand-1 (PD-L1), expressed by cancer and myeloid cells, suppress immune responses by engaging programmed-death-1 (PD-1) receptors on activated lymphocytes (*8, 9*). The PD-1/PD-L1 axis is a key mechanism of tumor-mediated immunosuppression, and its blockade has demonstrated remarkable clinical benefit across multiple solid tumors (*10–14*). Avelumab, a PD-L1 checkpoint inhibitor, is approved by the FDA to treat metastatic Merkel cell carcinoma, urothelial carcinoma, and renal cell carcinoma (*15–17*). Notably, meningiomas lie outside the blood-brain barrier and are therefore more accessible to peripheral immune cells (*18, 19*). PD-L1 is expressed by meningioma cells and tumor-infiltrating myeloid cells (*20, 21*), with expression levels correlating with higher tumor grade, recurrence after RT (*21, 22*), and enrichment of immunosuppressive cells in the tumor microenvironment (TME) and peripheral blood (*23*).

Evidence supporting checkpoint inhibition in meningioma remains limited. A case report described a patient with advanced lung cancer and an incidental intracranial meningioma who experienced a 24% reduction in meningioma volume following six months of PD-1 inhibitor therapy (*24*). In a retrospective analysis of 25 patients with incidental meningiomas receiving checkpoint inhibitors for other malignancies, 25% demonstrated tumor volume reduction (mean: −21%; SD: 6%) (*25*). However, three prospective single-arm phase II trials evaluating PD-1 inhibitors in RR-meningiomas reported only modest activity, with a combined partial response rate of 3% (2 of 65 patients) and PFS-6 ranging from 11% to 48% (*26–28*). Notably, one of the two partial responses occurred in a mismatch repair (MMR)-deficient tumor, a known predictor of response to PD-1 blockade and an established FDA-approved indication (*29–31*).

Given that meningiomas, in the absence of MMR deficiency, exhibit limited response to anti-PD-1 monotherapy, combination strategies may be required to enhance therapeutic efficacy. In a preclinical meningioma model, the combination of avelumab with high-affinity natural killer cells resulted in superior tumor control and survival compared to either agent alone (*32*). Growing preclinical data suggest that ionizing radiation can modulate TME to augment immunotherapy responses in poorly immunogenic solid tumors (*33–35*). Reported mechanisms include upregulation of major histocompatibility complex (MHC) class I and intercellular adhesion molecule-1 (ICAM-1) expression, secretion of proinflammatory cytokines such as CXCL16, and expansion of T-cell receptor (TCR) repertoire diversity among tumor-infiltrating lymphocytes (TILs) (*35–38*). Despite these preclinical findings, *in-vivo* validation of radiation-induced immunologic modulation of human meningiomas remain limited. To address this gap, we conducted a window-of-opportunity (WOO) clinical trial to evaluate the immunologic effects of combining neoadjuvant RT plus PD-L1 inhibitor followed by surgery in RR-meningiomas. This study also aimed to assess preliminary clinical efficacy and generate correlative insights into treatment-induced changes in tumor immunogenicity.

## RESULTS

### Patient Characteristics

Between Feb 2018 to Feb 2022, 11 patients were screened for the study, and 9 patients were enrolled and received neoadjuvant avelumab plus hypofractionated PBT, followed by surgery and adjuvant avelumab (study schema: **Fig. 1A**; CONSORT diagram: **fig. S1A**). Pre- and post-treatment tumor and blood specimens underwent multi-omic correlative analyses (**fig. S1B-C**): whole exome sequencing (WES), RNA sequencing (RNA-seq), multiplex immunofluorescence (MxIF), single-nucleus RNA sequencing (snRNA-seq), and flow cytometry (FC). The baseline patient characteristics are detailed in **Table 1**. Five patients had baseline archival tissue from initial surgery before RT (radiation-naïve), two patients (PT6&9) had baseline tissue from salvage surgery after prior RT (radiation-relapsed), and two patients (PT5&8) had baseline tissue from both initial surgery and salvage surgery. However, the baseline tissues from PT5 failed RNA-seq after multiple attempts, likely due to degraded tumor quality, but PT5’s baseline radiation-relapsed tissue was processed successfully for WES, while the baseline radiation-naïve tissue was processed successfully for MxIF. Based on WES, none of the patients had MMR deficiency, and all had low TMB as expected (**fig. S2A**). Meningiomas can be broadly classified into three molecular classes (MenG A-C) using DNA methylation, RNA-seq, or cytogenetic profiling, with MenG-C class (e.g. Hypermitotic meningiomas) associated with malignant behavior. Integrating multiple methods can also improve the accuracy of classification (*39*). Using both WES-based cytogenetic profiling (*40*) and RNA-seq method (*41, 42*), all the baseline and post-treatment tumors in this study were classified as MenG-C class (**fig. S2B-C**), confirming that all the RR-meningiomas in this study have malignant biology and that the molecular class probably does not change with treatment. Chen et al. have previously developed a 34-gene assay that can stratify meningiomas into three risk groups, which can predict PFS, OS, and RT responses (*43, 44*). RNA-seq data was used to classify the risk group of our baseline tumors, and 66% were classified as intermediate- or high-risk group (**Table 1**). Among the six patients with radiation-naïve baseline tumors, three were classified intermediate risk (PT2,3,7), and one each for low-risk (PT1), high-risk (PT4), and unknown (PT5, as RNA-seq failed). Two patients with radiation-relapsed baseline tumors were each classified as low (PT9) and intermediate risk (PT6). The patient with both radiation-naïve and radiation-relapsed baseline tumors (PT8) were classified as intermediate and high risk, respectively. In this cohort of RR-meningiomas, most patients were heavily pretreated, with majority of patients progressed after 2 surgeries, 2 courses of RT, and prior systemic therapy **(Table 1)**.

**Fig. 1.**
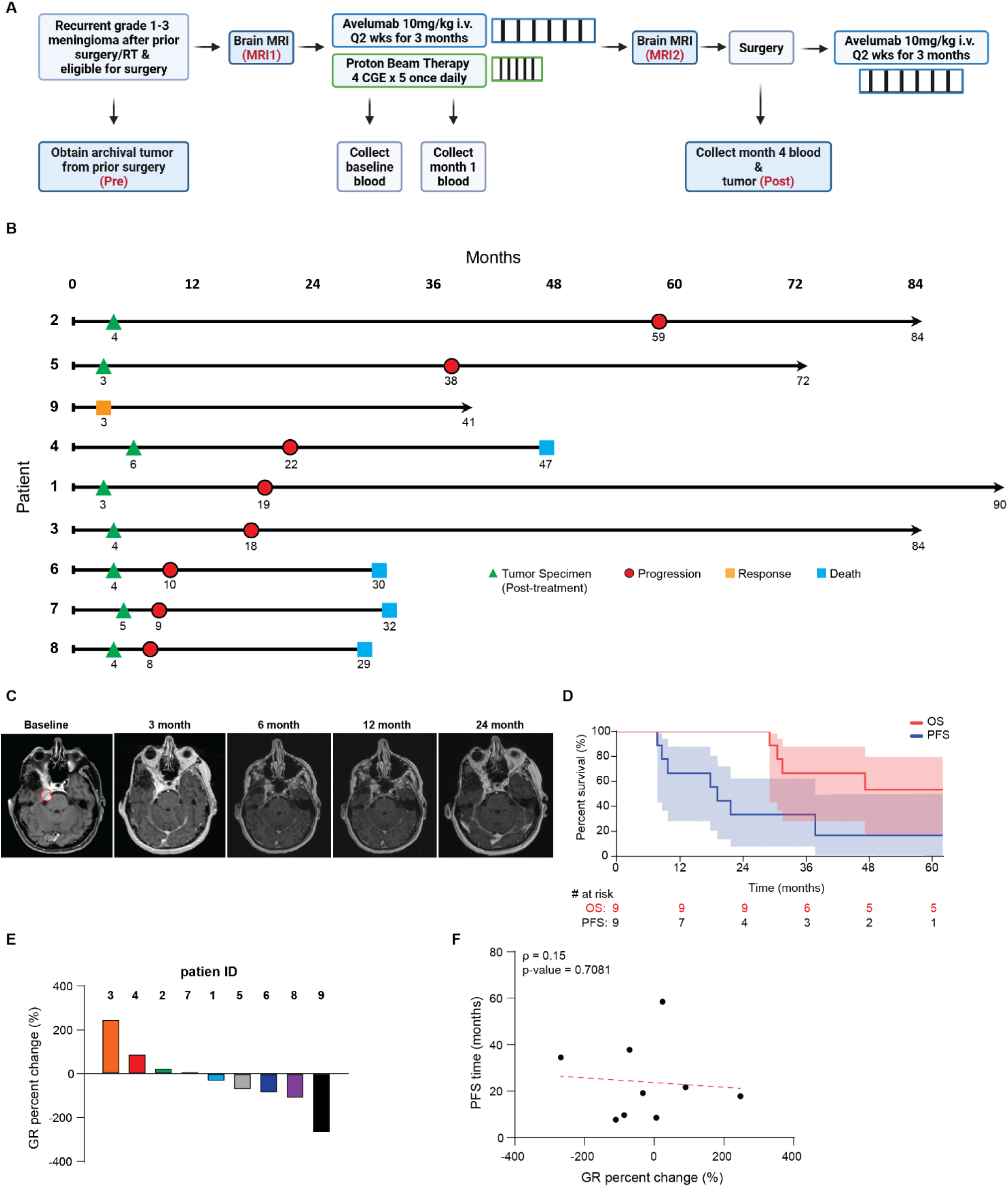
Radio-immunotherapy (avelumab and moderate-dose proton beam therapy) led to prolonged progression-free survival in a subset of patients with radiation-relapsed meningiomas. (A) Study schema. (B) Swimmer plot with time to event (months) from the start of study therapy for the nine evaluable patients. Each lane denotes individual patient’s clinical course and tumor response. (C) Radiographic changes of brain MRI during and after treatment for patient 9, who did not have planned surgery due to radiological response. (D) progression-free survival (PFS) and overall-survival (OS). Shaded areas represent 95% CI. (E) Growth rate (GR) percent change was calculated as the (post-treatment GR – pre-treatment GR)/pre-treatment GR × 100. Post-treatment GR was calculated as (tumor volume after neoadjuvant therapy – baseline tumor volume)/ time interval between MRIs x 12 months. Pre-treatment GR was calculated as (baseline tumor – prior tumor volume 5-10 months before study enrollment)/time interval between MRIs x 12 months. (F) Scatterplot showing the correlation between GR percent change and PFS. Correlation was evaluated using Spearman’s coefficient (ρ).

**Table 1:**
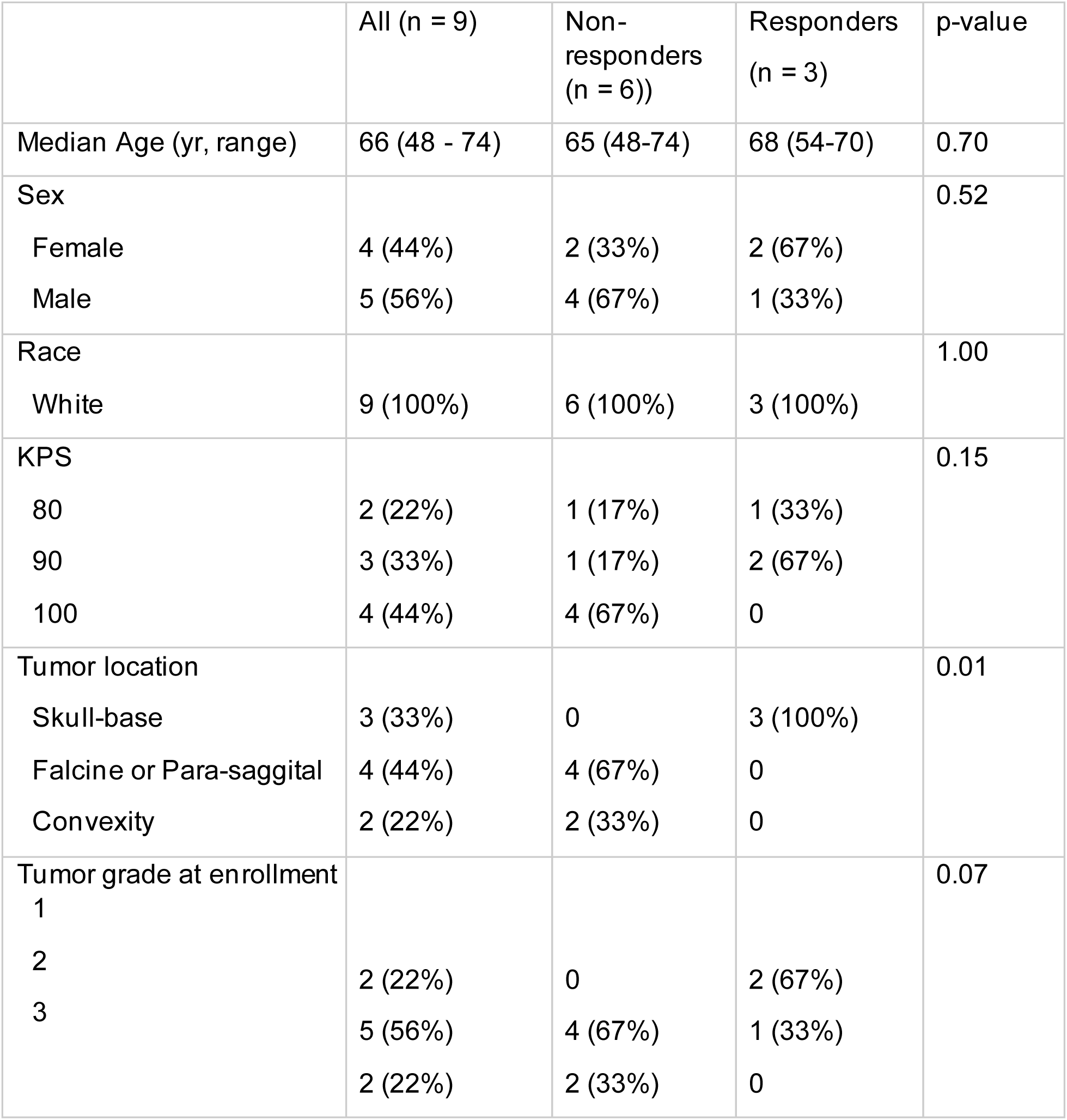
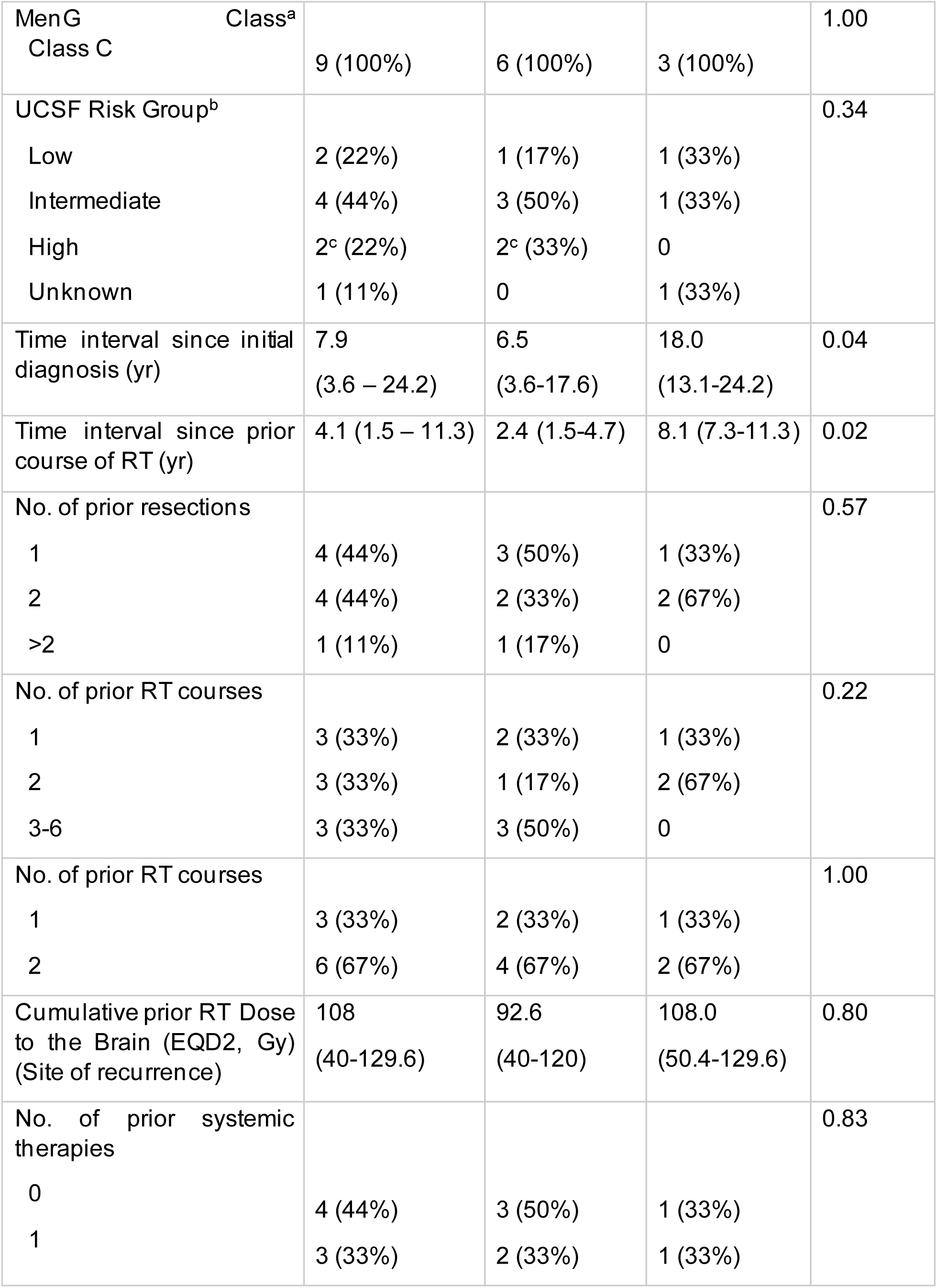
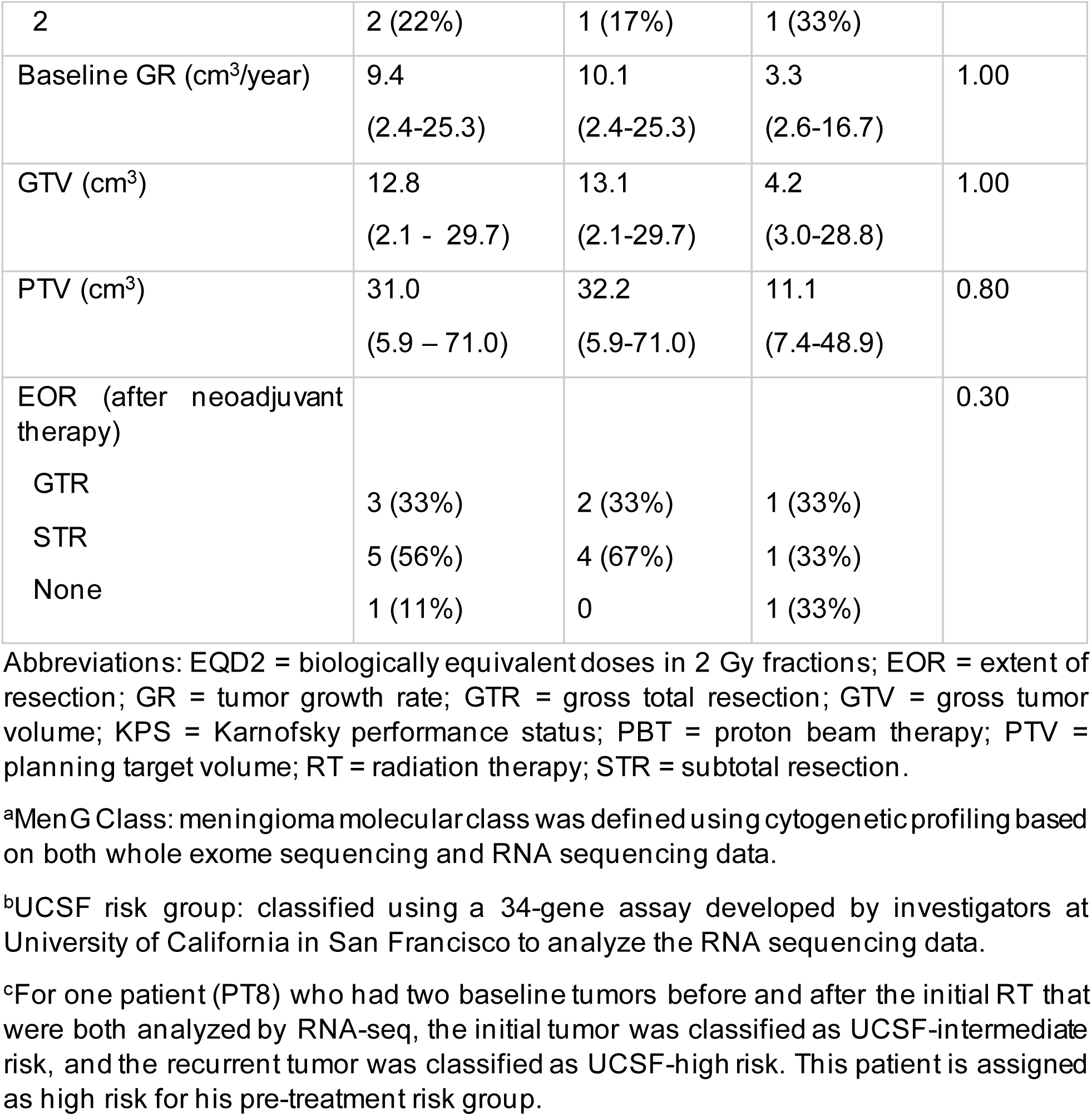
Patient and treatment characteristics.

### Safety and Clinical Outcomes

All patients completed neoadjuvant therapy, and all except one proceeded with surgery afterward. One patient forwent surgery due to dramatic radiological response. Two patients did not complete adjuvant avelumab due to immunotherapy-related diarrhea and infusion reaction, respectively. There was no dose-limiting toxicity (DLT), unexpected toxicity, or delayed radiation injury in the initial six-patient safety run-in or the overall cohort. Adverse events at least possibly related to study therapy are detailed in **Table 2**.

**Table 2:** Grade 2 or higher adverse events at least possibly related to study therapy.

|  | Grade 2 | Grade 3 | Total |
| --- | --- | --- | --- |
| Gastrointestinal |  |  |  |
| Diarrhea | 1 (11%) | 1 (11%) | 2 (22%) |
| General disorder |  |  |  |
| Fatigue | 1 (11%) |  | 1 (11%) |
| Infusion-related reaction | 1 (11%) | 1 (11%) | 1 (22%) |
| Investigations |  |  |  |
| Lymphopenia | 1 (11%) |  | 1 (11%) |
| Musculoskeletal |  |  |  |
| Generalized muscle weakness | 1 (11%) |  | 1 (11%) |
| Nervous system |  |  |  |
| Headache |  | 1 (11%) | 1 (11%) |
| Seizure | 1 (11%) |  | 1 (11%) |
| Sensory neuropathy | 1 (11%) |  | 1 (11%) |
| Trigeminal nerve disorder | 1 (11%) |  | 1 (11%) |
| Tremor | 1 (11%) |  | 1 (11%) |

As seen in **Fig. 1B**, three patients demonstrated prolonged PFS (58.5, 37.7, and 41.3+ months) including one patient with sustained radiological response who forwent surgery who is still in remission after 41.3 months (**Fig. 1C**). As seen in **Table 1**, three patients with prolonged PFS (hereafter referred as responders) had tumors at skull-base location and longer interval from the initial diagnosis and prior RT compared to the six patients with shorter PFS (hereafter referred as non-responders), while the other baseline characteristics and extent of resection post-neoadjuvant therapy were not significantly different between the two groups (**Table 1**).

After a median follow-up of 47.2 months (range: 29.0 – 89.7) and a minimum follow-up of 41.3 months for the last enrolled patient, eight patients have progressed, and four patients have passed after progression. As seen in **Fig. 1D**, the median PFS was 19.1 months (95% CI: 15.2-23.0), with one-year and two-year PFS rates of 67% and 33%, respectively. The median OS had not been reached, and the five-year OS was 53%. Five patients showed reduction of tumor growth rate (GR) after neoadjuvant therapy (as compared to tumor growth rate 5-10 months before study therapy), and two patients had stable GR. All three responders had either decreasing or stable ΔGR, but three non-responders also had decreasing ΔGR (**Fig. 1E**). ΔGR did not show significant correlation with PFS (**Fig. 1F**).

### TME Characterization by RNA-seq

Based on analysis of publicly available RNA-seq and microarray datasets of 735 predominantly treatment-naïve meningioma samples, we identified 4 microenvironment functional portrait (MFP) signatures: desert, myeloid/fibrotic, vascularized, and immune-enriched (**Fig. 2A**). Desert and myeloid/fibrotic have worse PFS compared to vascularized and immune-enriched MFP (**Fig. 2B**). RNA-seq analysis of our tumor samples showed an increase in T-cell and macrophage gene expression change of MFP in some post-treatment samples compared to pre-treatment samples, and some patients demonstrated transition from one MFP to a different MFP (**Fig. 2B-C**). Notably, two responders (PT2&5) exhibited immune-enriched phenotype post-treatment, while the third responder (PT9 who did not have surgery) had a pre-treatment MFP signature close to the immune-enriched type (**fig. S3A**). The pre-treatment tissue of PT5 yielded low quality RNA-seq data, so no pre-treatment MFP was determined. In contrast, none of the 6 non-responders showed transition to immune-enriched MFP post-treatment (**fig. S3B**). PT8 had two pre-treatment samples from two prior surgeries, and all three samples showed myeloid/fibrotic MFP (**fig. S3C**).

**Fig. 2.**
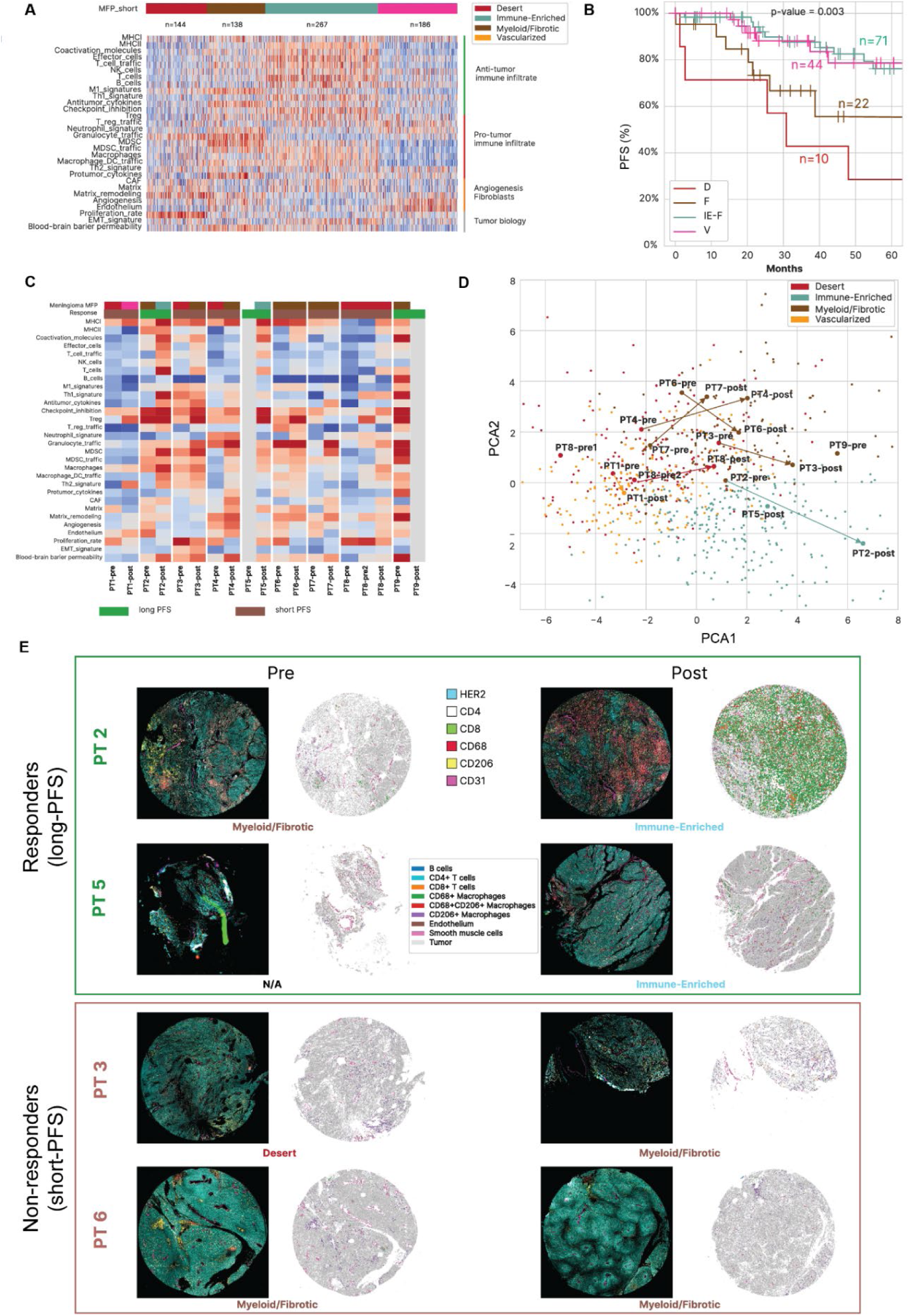
Radio-immunotherapy induced tumor microenvironment (TME) changes in radiation-relapsed meningiomas. (A) Heatmap depicting the differential gene expression of four distinct TME subtypes or Microenvironment Functional Portrait (MFP) signatures using a pooled cohort of meningiomas (n = 735) from publicly available RNA-seq and microarray datasets. (B) Progression-free survival (PFS) stratified by MFP signatures among a subset of 147 patients with available PFS data. (C) Heatmap depicting the differential gene expression of the pre- and post-treatment tumors of the 9 evaluable patients and the corresponding MFP signature derived from RNA-seq. Patient 5’s pre-treatment tumor samples failed RNA-seq, and patient 9 did not have post-treatment tissue. (D) Principal component analysis (PCA) plot depicting dynamic changes of MFP signatures after radio-immunotherapy. (E) Multiplex immunoflurorescence (MxIF) images of selected pre- and post-treatment tumors among responders (long PFS > 24 months) and non-responders (short PFS < 24 months).

### Characterization of T cells and macrophages on MxIF

To further investigate changes of immune cells in the TME, we selectively performed MxIF on 2 responders (PT2&5) and 2 non-responders (PT3&6). MxIF revealed a marked increase in T-cell and CD68^+^ macrophage infiltration in the responders post-treatment, a pattern not observed in the non-responders (**Fig. 2E**). Percentages of T cells and macrophages on MxIF also correlated well with reconstructed percentages of T cells and macrophages estimated from RNA-seq (**fig. S3D**). Particularly, the responders displayed increased infiltration of activated CD8^+^ T cells (Ki67^+^ or GZB^+^) as well as M1-phenotype macrophages (CD68^+^HLADR^+^ or CD68^+^Ki67^+^). In contrast, the pre- and post-treatment tissues from non-responders predominantly featured M2-phenotype (CD206^+^) macrophages (**Fig. 3A**). Community analysis of MxIF showed that the responders had tumor regions enriched with CD68^+^/CD206^−^ macrophages, while the non-responders had tumor regions enriched with CD206^+^ macrophages (**Fig. 3B**).

**Fig. 3.**
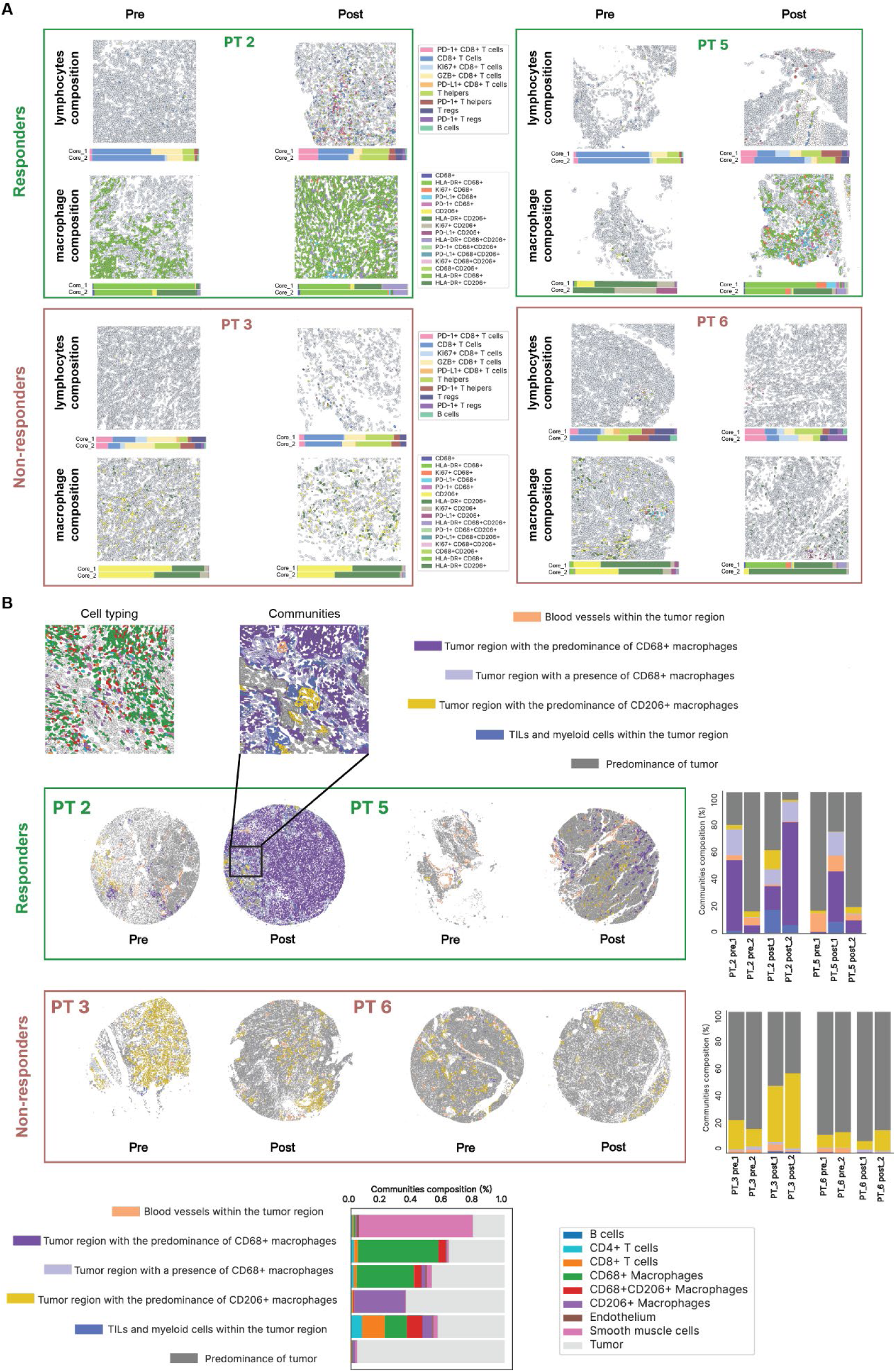
Differential immune cell infiltration between responders and non-responders after radio-immunotherapy for relapsed meningiomas. (A) Multiplex immunoflurorescence (MxIF) images of infiltrating lymphocytes and macrophages in the pre- and post-treatment of selected responders and non-responders. Horizontal color bar graphs below the representative image depict cellular composition from 2 separate cores for each tumor sample. (B) Community analysis depicting the six community cell clusters, as classified by dominant cell types and structural composition. Vertical color bar graphs depict cellular composition from two separate cores for each tumor sample.

### Macrophage Characterization using snRNA-seq

Given the striking difference in macrophage infiltration observed between responders and non-responders on MxIF, we next performed snRNA-seq on post-treatment snap-frozen tissues from the four patients who underwent MxIF. Macrophages accounted for ∼20% of all cells captured from the tissues (**Fig. 4A-B, fig. S4A–D**) and were further subdivided into 11 unsupervised clusters (**Fig. 4C, fig. S5A-B**). Notably, cluster 7 (C7) macrophages were specifically enriched in non-responders (**Fig. 4D**). To further characterize this population, we performed cell–cell communication analysis (**fig. S5C**). While no clear interactions between T cells and macrophages were detected (**fig. S5D**), C7 macrophages exhibited strong interactions with meningioma cells (**Fig. 4E**) through the fibronectin-1 (FN1) signaling pathway, uniquely serving as both sender and receiver (**Fig. 4F**). In comparison with other macrophage clusters, C7 macrophages demonstrate enriched expression of multiple FN1 receptors (**Fig. 4G**). These findings suggest that C7 macrophages may be stimulated by FN1 secreted from meningioma cells and contribute to the immunosuppressive TME in non-responders, which have been observed in other cancers (*45, 46*).

**Fig. 4.**
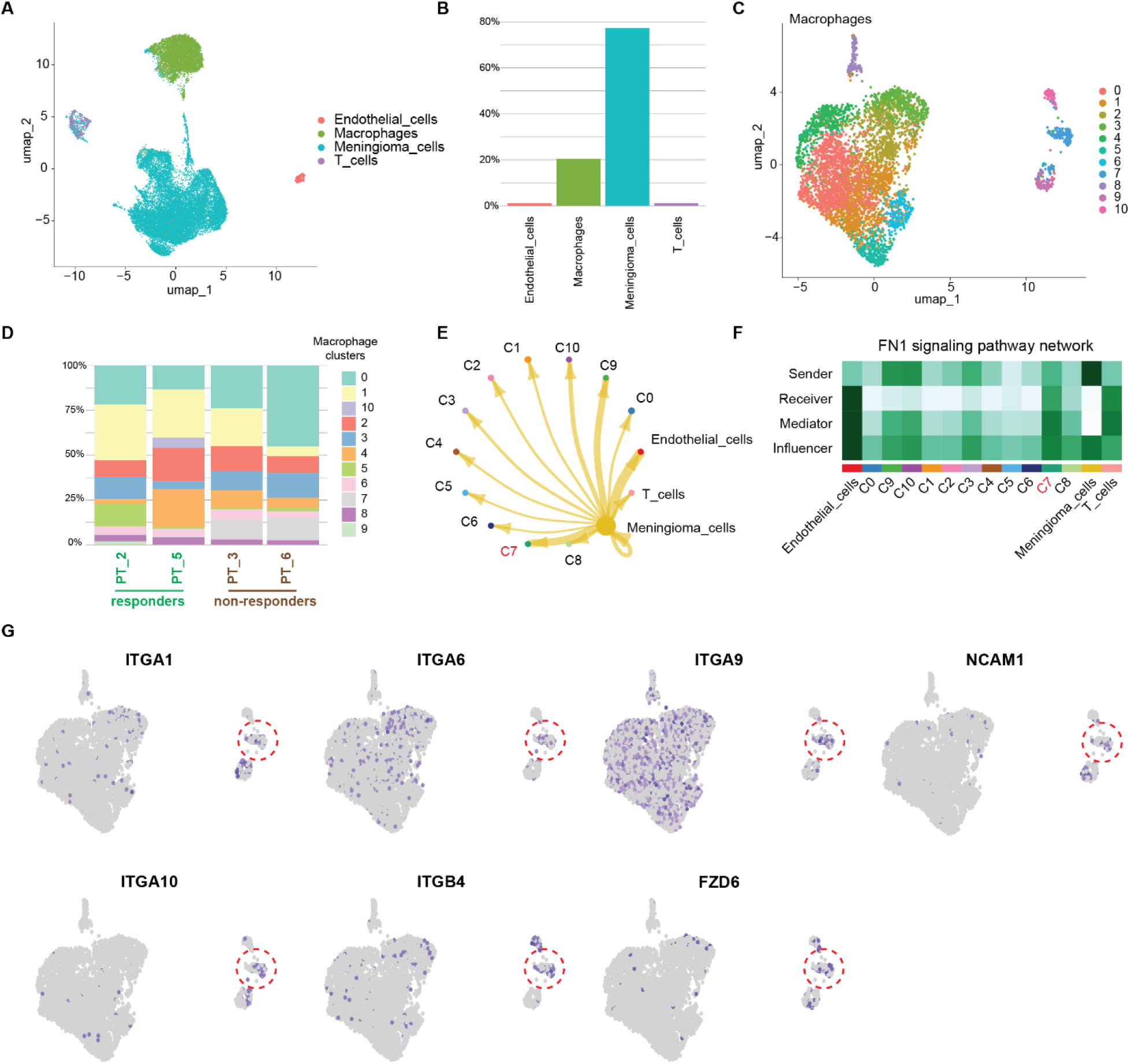
SnRNA-seq analysis on immune cells and macrophage subclusters of post-treatment patient tumor samples. **(A)** Uniform manifold approximation and projection (UMAP) plot of 25,983 cells, color-coded by associated cluster (each point represents a single cell). Cells were identified as four different major cell types using the expression of corresponding marker genes for each cell type. **(B)** Percentage of each identified cell type in the samples. **(C)** Macrophages were re-clustered into 11 subclusters. **(D)** Percentage of macrophage clusters of each patient tumor sample, separated by responders and non-responders. **(E)** Cell-cell interaction of tumor cells with macrophage clusters, endothelial cells, and T cells. The thickness of the lines represents the weighted strength of the interaction between the tumor cell and the corresponding cell population. **(F)** The strongest signaling pathway that establishes the interaction between cluster 7 macrophages and tumor cells. **(G)** Feature plots showing the expression of FN1 receptors. Cluster 7 macrophage population is outlined in red circles.

### Immune cell changes in peripheral blood on FC

The snRNA-seq analysis of post-treatment tumors showed the responders have higher proportion of infiltrating T cells (**Fig. 5A**) and dendritic cells (**Fig. 5B**) than non-responders. To further analyze the peripheral immune response, we compared FC data of PBMCs from three responders (PT2, 5, 9) and three non-responders (PT1, 3, 4), classifying samples into five immunophenotypes (G1-5) as previously described (*47*). In addition, we applied immunophenotype signature scores (ISSs), which quantified the similarity of each sample to the most representative instance of a given immunophenotype on a scale from 0 to 10 (**Fig. 5C-D**). The largest ISS value among all five ISS values from a patient’s blood sample was used to assign that sample to an immunophenotype. One month after initiating radio-immunotherapy, responders showed an increase in the G2-primed score, which is associated with expansion of CD4^+^ T cells, including differentiated central and transitional memory subsets (**Fig. 5C-E**). Responders also showed increased percentage of CD8^+^Ki67^+^ and CD8^+^Ki67^+^PD1^+^ T cells at month 1 and month 4 as compared to non-responders, which is consistent with the intratumoral T-cell infiltration in **Fig. 3A**. In contrast, no difference was observed for CD4^+^Ki67^+^ and CD4^+^Ki67^+^PD1^+^ T cells (**Fig. 5E-H**). These data suggest that responders may have a different peripheral immune response compared to non-responders.

**Fig. 5.**
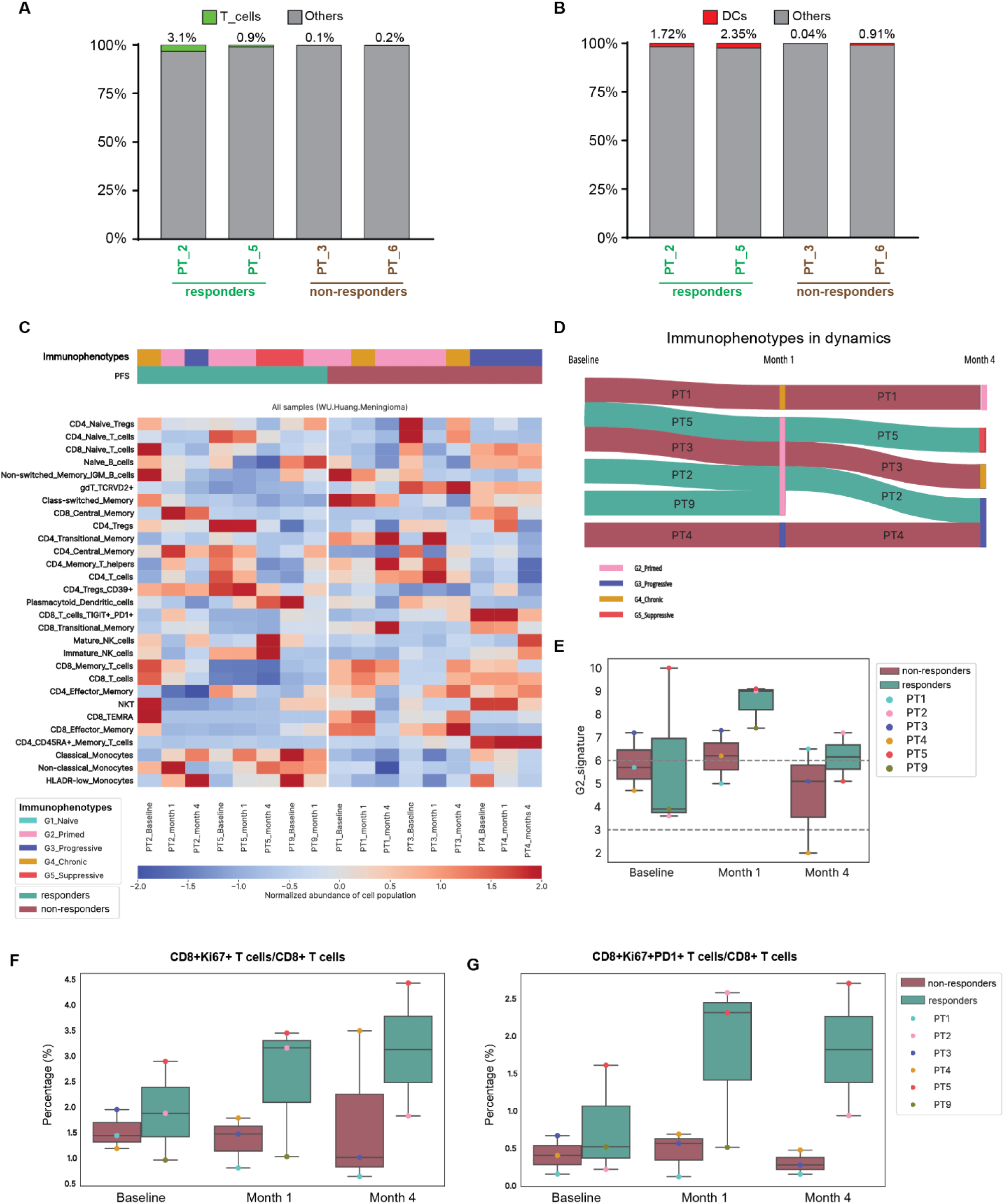
Tumor-infiltrating immune cells and peripheral immune cell changes after radio-immunotherapy. Percentage of **(A)** T cells and **(B)** dendritic cells of each patient tumor sample as identified via snRNA-seq, separated by responders and non-responders. (C) Spectral clustering analysis applied to normalized flow cytometry percentages for selected pre- and on-treatment peripheral blood samples of 6 patients and the corresponding blood immunophenotypes. (D) Sankey diagram showing the changes of each patient’s blood immunophenotypes during treatment. (E) Box plots comparing G2-primed score of blood samples between responders and non-responders before and during treatment. G2-primed score quantifies the similarity of each sample to the G2-primed immunophenotype on a scale from 0 to 10, with 10 representing perfect similarity. (F-G) Box plot comparing proportion of CD8^+^Ki67^+^ T cells or CD8^+^Ki67^+^PD1^+^ T cells among all CD8^+^ T cells between responders and non-responders before and during treatment.

## DISCUSSION

This study demonstrated that a subset of RR-meningiomas can mount an immunological response and experience prolonged PFS following PD-L1 inhibition combined with short-course hypofractionated RT, supporting this approach as a promising strategy for further evaluation in larger-scale trials. However, the lack of response observed in the majority of RR-meningiomas underscores the need for predictive biomarkers to guide treatment selection. Correlative analyses from this trial suggest that pre-treatment myeloid cell signature within the TME and peripheral primed T-cell signature one-month post-treatment may serve as potential predictors of resistance and response.

This study demonstrated that 33% of unselected RR-meningiomas developed a robust immunological response and prolonged PFS following neoadjuvant PBT (4 CGE x 5 fractions) combined with a PD-L1 inhibitor. To our knowledge, this is the first clinical study to clearly demonstrate an on-target immune response to the combination of RT with PD-1/PD-L1 blockade in RR-meningiomas, providing clinical proof of concept to preclinical studies that suggest radio-immunotherapy may be beneficial in poorly immunogenic tumors (*33–35*). This study answers the call to develop more WOO studies in Neuro-Oncology to evaluate the biological plausibility of a novel therapeutic strategy in clinical setting before embarking in large-scale clinical trials (*48, 49*). A multi-institutional phase I/II study conducted through the NCI Experimental Therapeutics Clinical Trials Network (ETCTN-10186) is currently evaluating hypofractionated radiosurgery (8 Gy x 3 fractions) combined with PD-1 inhibition (nivolumab) with or without cytotoxic T-lymphocyte associated protein 4 (CTLA-4) blockade (ipilimumab) in patients with high-grade RR-meningiomas (NCT03604978). Interim results from the trial suggest both radio-immunotherapy regimens were well tolerated and produced radiological responses (*50*). The study recently completed accrual, totaling 6 patients with radiosurgery plus nivolumab and 24 patients with radiosurgery plus nivolumab and ipilimumab. The final results of ETCTN-10186 will provide important complementary data regarding the clinical efficacy of definitive radio-immunotherapy strategy in RR-meningiomas.

Selection of the primary endpoint will be critical for future large-scale trials evaluating radio-immunotherapy in RR-meningiomas. While the WOO design in this study enabled direct assessment of immunological response as the primary endpoint, such a strategy is not feasible for larger, multi-institutional randomized trials that would be difficult to mandate surgical sampling. Our findings support the use of PFS (as determined by the well-established iRANO criteria) as the primary endpoint in future trials, whereas early radiographic tumor growth rate changes after neoadjuvant radio-immunotherapy (compared to tumor growth rate prior to study therapy) did not correlate with PFS or immune response (**Fig. 1E-F**). Since meningiomas rarely undergo significant shrinkage with systemic therapy, changes in growth rate have been proposed as an early radiographic biomarker of treatment response (*51*). However, our results suggest this metric may not be reliable in the context of combination regimens that include RT, as RT can induce tumor shrinkage independently of immune activation. This finding has important implications in clinical trial design. Reliance on radiologic growth rate or volumetric response as surrogate endpoint may introduce complexity and misinterpret treatment efficacy, particularly in large, multi-center studies. Standardized and clinically meaningful endpoints such as PFS will be more practical and interpretable across diverse clinical settings.

The finding that 67% of patients in this study did not develop an immunological response to radio-immunotherapy underscores the heterogeneity of TME in meningiomas. This variability may explain the differing radiological responses observed in prior studies of single-agentanti-PD-1 blockade in prior studies: 25% with volume reduction in treatment-naïve incidental meningiomas (*25*) versus 3% response rate in RR-meningiomas across three single-arm phase II trials (*26–28*). Patients with RR-meningiomas enrolled in the prior phase II trials likely represented a heavily pretreated population enriched for biologically aggressive tumors with a more immunosuppressive TME. While our study also focused on RR-meningiomas, the requirement for candidacy for both re-irradiation and surgical resection and allowance of grade 1 tumor may have selected for a subset of tumors more amenable to immune modulation, as reflected by the longer interval since prior RT among responders compared to non-responders (**Table 1**). Analysis of publicly available RNA-seq and microarray datasets of predominantly treatment-naïve meningiomas further support this concept, showing that a subset of meningiomas exhibiting immune-enriched TME signature have superior PFS compared to immune-desert or myeloid/fibrotic subtypes (**Fig. 2A-B**). These immune-enriched meningiomas may correspond to the incidental meningiomas that responded to checkpoint inhibitor alone in prior retrospective reports (*24, 25*). Importantly, none of the RR-meningiomas in our study exhibited an immune-enriched TME before study therapy, which may partially explain their relapse and treatment resistance (**Fig. 2C-D**). However, our data suggest that a subset of myeloid/fibrotic meningiomas may acquire immune-enriched features when treated with the combination of RT and PD-L1 blockade, providing a rationale for further exploration of dynamic TME modulation as both a biomarker and therapeutic strategy.

Predictive biomarkers that enable identification of likely responders before treatment initiation are critical to optimize the therapeutic ratio of radio-immunotherapy. This study suggests that myeloid cells within the TME may play a key role in mediating response, as responders exhibited marked post-treatment changes in myeloid populations (**Fig. 3**). Interestingly, all responders had skull-base meningiomas, which are known to have distinct immune microenvironment compared to non-skull-base meningiomas, including closer proximity to meningeal lymphatic vessels and increased infiltration of Tregs and M2-phenotype macrophages (*52–54*). Our snRNA-seq revealed heterogeneous myeloid cell states and suggested that FN1-associated macrophages (C7 macrophages) may contribute to resistance to radio-immunotherapy (**Fig. 5D-G**). FN1 has been shown to polarize tumor-associated macrophage (TAM) toward immunosuppressive state, and FN1-associated TAM has been correlated with immunotherapy resistance in multiple cancer types, including breast cancer, gastric cancer, and glioma (*55–58*). Although the limited sample size of our study precludes the establishment of specific predictive myeloid markers, the multi-omic correlative findings highlight the potential of myeloid cell composition and skull-base tumor location as predictive biomarkers for radio-immunotherapy response.

This study also suggests that immunophenotyping of peripheral blood one month after initiating PD-L1 blockade may serve as a promising early predictive biomarker of immunologic response. This has important clinical implications, particularly given that only a subset of patients may derive benefit from immunotherapy, so early identification of non-responders may enable timely discontinuation of ineffective treatment. Dyikanov et al. previously developed a multiparameter FC-based immunoprofiling platform that classifies peripheral immune cells into five conserved immunophenotypes. Using this framework, they generated a continuous scoring system (ISSs) capable of characterizing individual patient samples with robust performance, even in small cohorts. They previously applied this approach to predict immunotherapy response using blood samples from a clinical trial of 35 patients with head and neck squamous cell carcinoma treated with anti-PD-1 blockade. They observed on-treatment samples from responders were predominantly classified as the G2-primed immunophenotype, and G2-primed ISSs of on-treatment samples distinguished responders from non-responders with 76% accuracy (*47*). When we applied this same method to our cohort, G2-primed ISSs were significantly higher in responders than non-responders one month after treatment initiation (**Fig. 4C**), supporting its potential utility to monitor immunotherapy response in RR-meningiomas. In a separate study of 29 patients with lung cancer treated with PD-1 inhibition, Kamphorst et al. reported that an early increase of CD8^+^Ki67^+^PD1^+^ T cells one month after treatment was associated with improved clinical outcomes: 80% of responders exhibited an early increase, while 70% of non-responders had no or delayed increase (*59*). Similarly, in a study of 29 patients with melanoma treated with PD-1 inhibition, Huang et al. found that a higher ratio of CD8^+^Ki67^+^PD1^+^ T-cell increase relative to tumor burden after initiating treatment correlated with improved outcomes (*60*). In our study, responders also exhibited a higher proportion of CD8^+^Ki67^+^PD1^+^ T cells one month after initiating immunotherapy compared to non-responders (**Fig. 4F**). Prospective validation in larger cohorts will be necessary to determine whether G2-primed ISS or CD8^+^Ki67^+^PD1^+^ T-cell proportion provides more reliable predictive performance. The observed differential peripheral immune responses and the corresponding intratumoral T-cell infiltration in the responders also suggest that systemic immune status, rather than the TME alone, may play a key role in driving immunotherapy response, warranting further investigation.

The major limitations of this study include its small sample size, lack of immediate pre-study treatment tumor tissue, and absence of a randomized control group. Although the study initially aimed to enroll 12 patients, it was terminated early after accruing 9 patients due to withdrawal of drug support from Pfizer and Merck KGaA. However, the optimal sample size for WOO studies design is not well established, and prior neuro-oncology WOO studies evaluating biological endpoints typically have a sample size of 3-6 patients per treatment arm (*48*). Since the trial was designed with surgical resection following neoadjuvant therapy, baseline tumor tissue immediately prior to treatment was not available. Mandating a pre-treatment biopsy was deemed not feasible, so archival specimens from prior surgeries were used as baseline controls. Most baseline archival samples that were successfully sequenced by RNA-seq (n=6) came from the initial surgery prior to RT, while three samples were collected from salvage surgery following RT. One patient had archival tissue from both time points, and RNA-seq of both samples showed no changes in TME phenotype before and after RT (**fig. S3C**). Notably, none of the three radiation-relapsed archival tissue exhibited immune-enriched MFP signature, suggesting RT alone is unlikely to induce immune enrichment in the TME. Our correlative analyses were further limited by technical issue with RNA-seq in one responder (PT5), and by the absence of post-treatment tissue in another responder (PT9, who opted out of surgery due to radiological response). Despite these constraints, the multi-omic approach of our correlative analyses enabled complementary insights into the immune responses elicited by the combination of RT and PD-L1 blockade, so we were able to extract maximum information from patients treated in this small study. As a single-arm phase I study, this trial cannot exclude the possibility that the observed immunological responses in responders could not have resulted from either RT alone or PD-L1 inhibitor alone. However, all three responders had previously progressed after receiving much higher-dose RT and lacked baseline immune-enriched signature by RNA-seq and/or MxIF. These findings make it unlikely that either low-dose RT or PD-L1 monotherapy would have triggered the robust immune responses and durable PFS observed. Nonetheless, larger and randomized studies will be necessary to confirm these promising findings and validate the clinical benefit of this radio-immunotherapy strategy for RR-meningiomas.

## MATERIALS AND METHODS

### Study design

This single-institution, single-arm, open-label, phase 1 WOO study aimed to investigate the on-target immunologic effects of combining RT with PD-L1 inhibition in RR-meningiomas. The primary objective was to evaluate changes in immune cell populations, specifically T cells and myeloid cells, within the TME three months after radio-immunotherapy. Secondary objectives included assessment of safety, radiographic tumor growth, and clinical efficacy as measured by progression-free survival (PFS) and overall survival (OS). Exploratory objectives included identification of tumor-associated genomic signatures or peripheral blood biomarkers predictive of response to radio-immunotherapy. The study was designed with an empirical sample size of 12 patients, selected based on feasibility and recommended sample size for exploratory pilot studies (*61*). However, the study terminated early after enrollment of 9 patients due to slow accrual during the COVID-19 pandemic and changes in collaboration between Pfizer and Merck KGaA to co-develop avelumab. An interim safety analysis was planned after the first 6 patients completed at least 6 months of follow-up. Prespecified safety thresholds were: <33% of patients experiencing acute dose-limiting toxicity (DLT) or delayed radiation injury. DLT was defined as grade 3 or higher treatment-related adverse events that were at least possibly related to either avelumab or PBT within 12 weeks of the start of therapy, excluding grade 3 infusion-related reaction resolving within 6 h and controlled with medical management, transient grade 3 influenza-like symptoms or pyrexia controlled with medical management, grade 3 fatigue, grade 3 nausea/vomiting, grade 3 diarrhea, grade 3 skin toxicity, or grade 3 elevated liver enzymes that resolved to grade 1 within 7 days after initiation of medical management. Delayed radiation injury was defined as grade 3 or higher radiation necrosis or symptomatic edema at any time after the initial 12-week DLT period. The study protocol was approved by the institutional review board and conducted in accordance with the Declaration of Helsinki and Good Clinical Practice guidelines. An Investigational New Drug (IND) application was approved by the U.S. Food and Drug Administration. All patients provided written informed consent prior to enrollment. The trial was registered with Clinicaltrials.gov (NCT03267836). Avelumab, the PD-L1 inhibitor used in this study, was provided by Pfizer and Merck KGaA.

### Eligibility

Patients were eligible if they had a clinical diagnosis of recurrent WHO grade 1-3 meningioma following prior surgery and RT, were ≥ 18 years old, had a Karnofsky performance status ≥ 60, and had archival tumor tissue available for baseline comparison. Additional eligibility criteria included suitability for repeat surgery after a 3-month neoadjuvant period, a dexamethasone dose ≤ 4mg daily, and adequate hematological, renal, and hepatic function. Prior RT could include external beam RT, radiosurgery, or both; there was no limit on the number or cumulative dose of prior RT courses. Key exclusion criteria included: prior therapy targeting PD-1 or PD-L1, active infection requiring systemic therapy, known infection related to human immunodeficiency virus, hepatitis B virus, or hepatitis C virus, concurrent use of systemic immunosuppressive medication or other investigational agents, prior hypersensitivity to checkpoint inhibitors, history of organ transplant or auto-immune disease, pregnancy, or other medical illness contraindicating immunotherapy.

### Treatment

Patients received neoadjuvant avelumab (10mg/kg IV) every two weeks for 3 months, for a total of six doses. PBT was administered concurrently at 4 cobalt gray equivalents (CGE) per day for 5 daily fractions, starting within three days of the first avelumab dose. This hypofractionated regimen was selected based on preclinical and clinical evidence suggesting that regimens within 3-5 fractions and > 2 Gy per fraction are more effective at inducing immunostimulatory response than conventional fractionation (1.8-2 Gy per fraction) or a single fraction of high dose RT (*34, 62*). A prospective phase II study also demonstrated the safety and efficacy of hypofractionated RT (5 Gy x 5 fractions) for large or optic nerve-adjacent, radiation-naïve meningiomas (*63*). Additional factors influencing the dose selection included uncertainty about the higher relative biological effectiveness (RBE) of PBT, desire to include large recurrent meningiomas with significant prior radiation exposure, concurrent avelumab administration, and planned surgery following neoadjuvant therapy. As a result, the RT dose was reduced by 20% relative to the 5-fraction hypofractionated RT regimen. For RT planning, gross tumor volume (GTV) was defined as contrast-enhancing tumor on the simulation MRI, excluding prior surgical cavities or dural tails. Simulation CT was also used to assist tumor delineation. Planning target volume (PTV) was defined as GTV plus 0.3 cm geometric margin. If the proton machine was unavailable due to mechanical issues, photon-based intensity-modulated RT was permitted as a substitute. All RT was required to be completed within 16 calendar days. Dosimetric constraints are detailed in **Supplemental Table S1**. All target contours and plans were centrally reviewed by the study principal investigator (JH) for compliance. After 3 months of neoadjuvant avelumab, patients underwent restaging MRI followed by planned surgical resection. Postoperatively, patient received an additional 3 months of adjuvant avelumab.

### Biospecimen Collection

Prior to study enrollment, archival formalin fixed paraffin-embedded (FFPE) tumor tissues from previous surgeries were obtained to serve as the pre-treatment baseline. The baseline tissue could originate from either the initial resection before any RT or salvage surgery following RT. After 3 months of neoadjuvant therapy, patients underwent surgical resection, and tumor specimens were divided: a portion was snap-frozen and cryopreserved, while the remainder was processed as FFPE tissue. Peripheral blood samples were collected at baseline, month 1 during neoadjuvant therapy, and before surgical resection (approximately month 4). Blood was processed to isolate peripheral blood mononuclear cells (PBMCs) and plasma, which were cryopreserved.

### Safety, Efficacy, and Radiographic Assessment

Patients were seen every 2 weeks during avelumab, and again at 3 and 6 months after completion to monitor for adverse events. Following treatment, brain MRI was performed every 3-6 months until progression. Tumor response and progression were assessed using the iRANO criteria (*64*). To calculate radiographic growth kinetics, a historical MRI (median 7.9 months before the baseline planning MRI; range 5.2 - 10.7 months) and the post-treatment MRI (median 3.4 months after neoadjuvant therapy; range 2.8 - 4.7 months) were imported into the RT planning software and co-registered with the baseline MRI. Enhancing tumor volumes on the historical MRI (GTV_historical_) and post-treatment MRI (GTV_post_) were manually contoured by the study PI. Pre-treatment GR = change in volume (cm^3^) from GTV_historical_ to GTV, normalized over 12 months; post-treatment GR = change in volume (cm^3^) from GTV to GTV_post_, normalized over 12 months. The percent change of GR (ΔGR) = (post-treatment GR – pre-treatment GR)/pre-treatment GR.

### Whole exome sequencing (WES) and RNA Sequencing (RNA-seq)

Tumor DNA and RNA were extracted from FFPE tissues using the Qiagen FFPE AllPrep DNA/RNA Kit. Normal DNA was extracted from blood using the Qiagen QIAamp DNA Blood Mini Kit. DNA Libraries were prepared using the Agilent SureSelect XT HS2 DNA Kit and hybridized with the Agilent SureSelect Human All Exon V7 capture panel. RNA Libraries were prepared using the Agilent SureSelect XT HS2 RNA Kit and hybridized with the Agilent SureSelect Human All Exon V7+UTR panel. Sequencing was performed on an Illumina NovaSeq 6000 system. Quality control of raw reads were conducted using FastQC v0.11.9, FastQ Screen v0.14.0, and MultiQC v1.4.

For WES, low quality reads were filtered with FilterByTile/BBMap v37.9075, then aligned to the GRCh38 reference genome (GRCh38.d1.vd1 assembly) using BWA v0.7.17.76. Duplicate reads were removed using Picard MarkDuplicates v2.6.0. Indels were realigned and recalibrated using IndelRealigner and BaseRecalibrator (GATK v4.1.2.0). Germline and somatic single-nucleotide variants, insertions, and deletions were detected using Strelka v2.9.10 and annotated using Variant Effect Predictor v92.1. Copy number alterations were assessed using a customized version of Sequenza v2.1.2. Microsatellite status was evaluated by MSI sensor v0.6. Tumor mutation burden (TMB) was derived from WES to quantify mutations per megabase (Mb) in the ∼30 Mb of coding regions of the genome.

RNA-seq reads were pseudo-aligned to GENCODE v23 transcripts using Kallisto v0.42.4 with default settings. Protein-coding genes, immunoglobulin heavy and light (kappa and lambda) chain transcripts, and TCR-related transcripts were retained. Noncoding RNA, histone genes, and mitochondrial transcripts were removed, yielding in 20,062 protein-coding genes. Gene expression was quantified as the summed Transcripts Per Million (TPM) values across transcripts and log2-transformed. Gene fusions were detected using STAR-Fusion v.1.8.1. Reconstructed percentages of T cells, macrophages, B cells derived from RNA-seq data were determined using Kassandra algorithm pre-trained on artificial transcriptomes (*65*).

### Tumor Microenvironment Functional Portrait (MFP)

We constructed a Microenvironment Functional Portrait (MFP) signature using a pooled cohort of meningioma patients from publicly available RNA-seq and microarray datasets (**supplemental Table S2**), following previously described method (*66*). Immune and stromal activities were inferred by single-sample gene set enrichment analysis (ssGSEA) across curated gene sets representing key TME components, including immune cell populations (e.g., macrophages, tumor-infiltrating lymphocytes), non-cellular factors (e.g., immunosuppressive cytokines, extracellular matrix), and malignant cell processes (e.g., proliferation) (*67, 68*). Signature activity scores were median-scaled within each dataset to minimize batch effects across platforms. Scaled scores were subjected to Leiden clustering, which identified four distinct TME clusters or MFP types: desert, immune-enriched, myeloid/fibrotic, and vascularized. Clustering parameters were optimized using the Calinski-Harabasz index, Silhouette score, and Davies-Bouldin index. RNA-seq data from this study cohort were subsequently classified into these MFP types using the same pipeline (*66*).

### Multiplex immunofluorescence (MxIF)

FFPE blocks from pre- and post-treatment tumor samples of 4 patients (2 non-responders and 2 responders) were processed for MxIF. Sections were stained using hematoxylin and eosin (H&E) and reviewed by a board-certified pathologist to assess for tumor purity and tissue quality. Two representative tumor regions (2 mm in diameter) were selected from each sample, resulting in 16 cores. These donor cores, along with tonsil and breast tissues controls, were arrayed into a 4 x 5 tissue microarray (TMA). Paraffin sections (4 µm) were stained with a 25-antibody panel plus DAPI (**Supplemental Table S3**) and imaged following the manufacturer’s protocol (Akoya Biosciences). Images were acquired at 20x using a 0.70 NA objective.

As a part of preprocessing, artifact detection was performed using a semantic segmentation model with U-Net++ architecture. Cell segmentation was performed using a U-Net-based neural network. Of the 26 stained markers, DAPI marked nuclei and NaK ATPase labeled membranes. The segmentation model was pretrained on a manually annotated internal dataset with three channels: nuclear, membrane, and a composite membrane/cytoplasmic marker channel. Each cell was uniquely tagged with a cell ID and spatial coordinates, and marker expression was quantified as mean intensity and area of expression.

After quality control by a pathologist, usable markers were selected based on intensity thresholds to exclude background staining. Each cell was annotated based on marker expression within its contour. Cell typing was then performed using Leiden clustering, identifying 9 major cell populations (**Supplemental Table S4**).

Subtyping of lymphocytes, macrophages, and tumor cells was performed based on additional marker expression. Key markers included Ki67, PD-1, PD-L1, and Granzyme B for lymphocytes; HER2, Ki67, PR, and Fibronectin for tumor cells; and HLA-DR, Ki67, PD-1, and PD-L1 for macrophages. Both mean intensity and area were used to define subtypes, and a gating approach was applied when signal intensity was low. Subtype identification was visually validated by overlaying cell masks on the original MxIF image, colored by subtype.

Community analysis was performed using a graph of cell centroids, incorporating nearby structural masks. Graphs were constructed with Delaunay triangulation, removing edges longer than 200 μm. Each node contained metadata on cell type, mean neighbor distance, and structure mask percentages (macrophages, tumor, endothelium) within a 65 μm radius—a validated spatial threshold for community analysis. A Graph Auto-Encoder was trained for 150 epochs to generate node embeddings. Encoded vectors (length = 32) were used for unsupervised clustering. The optimal number of clusters was estimated using the Calinski-Harabasz and Davies-Bouldin indices, resulting in six final community clusters. Clusters were classified by dominant cell type and structural composition (**Supplemental Table S5**). Spatial localization of these communities enabled analysis of their distribution relative to blood vessels. KDTree was used to calculate the distance (μm) of each community to the nearest vessel, allowing comparison between patients.

### Flow Cytometry (FC)

PBMCs were blocked with 300 μL of blocking buffer on ice for 10 minutes. Cells were then washed in PBS, centrifuged at 300 × g for 3 minutes, and stained with 50 μL of Ghost Viability Dye (0.25% v/v; 1:400, Tonbo) for 10 minutes at room temperature. After viability staining, cells were washed and extracellularly stained using four custom antibody panels (**Supplemental Table S6**) for 20 minutes at RT. Following staining, cells were fixed in 1% paraformaldehyde (Cytofix/Cytoperm, BD Biosciences) for 10 minutes at 4°C. For intracellular staining (A5P4 panel), cells were incubated with Foxp3 Fixation/Permeabilization buffer for 10 minutes at RT, washed twice with 1× permeabilization buffer, and stained with intracellular antibody cocktail (20 μL) overnight at 4°C. All samples were resuspended in 100 μL of acquisition buffer prior to analysis. Stained samples were acquired on a BD FACSymphony™ A5 Cell Analyzer. Compensation matrices were generated in FACSDiva software using single-stained controls. Raw .fcs files were transformed using the arcsinh function with a cofactor of 190 for marker channels. Size channels were scaled by dividing by 100,000 to align magnitudes. Cell populations were annotated through sequential manual labeling using FlowSOM clustering, two-dimensional scatter plots based on marker distribution, singlet gating via FSC-A vs FSC-H plots. To calculate cell frequencies across panels, results were normalized to a leading panel using reference population counts (T-cells or NK-cells + T-cells). Data processing was performed using Python 3.9 on the JupyterHub platform.

### Single Nucleus Suspension Preparation

Single-nucleus suspensions were prepared using the protocol adapted from the 10X Genomics Nuclei Isolation Protocol. Specifically, ∼20 mg of snap-frozen tumor tissue was transferred into a pre-chilled 1.5-mL microcentrifuge tube and chopped into small pieces using sterile scissors. The tissue was resuspended in 300 µL of NP40 Lysis Buffer (10 mM Tris-HCl, pH 7.4; 10 mM NaCl; 3 mM MgCl₂; 0.1% NP40; 1 mM DTT; 1 U/µL RNase inhibitor) and homogenized on ice 15 times using a pellet pestle manually. Following homogenization, 1 mL of additional NP40 Lysis Buffer was added, and the mixture was incubated on ice for 5 minutes. The lysate was filtered through a 70-µm cell strainer into a 2-mL microcentrifuge tube and centrifuged at 500 × g for 5 minutes at 4°C. The supernatant was carefully removed, and the pellet was resuspended in 1 mL of PBS containing 1% BSA and 1 U/µL RNase inhibitor. The suspension was incubated on ice for 5 minutes, gently pipette-mixed, and centrifuged again at 500 × g for 5 minutes at 4°C. The resulting pellet was resuspended in 100 µL of Lysis Buffer (1 mM Tris-HCl, pH 7.4; 1 mM NaCl; 0.3 mM MgCl₂; 0.1% BSA; 0.01% Tween-20; 0.1 mM DTT; 0.1 U/µL RNase inhibitor) and gently mixed by pipetting five times. The sample was incubated on ice for 2 minutes, followed by the addition of 1 mL Wash Buffer and another 5 times of pipette mixing. The mixture was centrifuged at 500 × g for 5 minutes at 4°C, and the supernatant was carefully removed without disturbing the nuclei pellet. Nuclei concentration was determined using a Countess II FL Automated Cell Counter (Thermo Fisher Scientific). Based on the nuclei count, samples were resuspended in an appropriate volume of chilled Diluted Nuclei Buffer (1X Nuclei Buffer (10x Genomics); 1 mM DTT; 1 U/µL RNase inhibitor) and proceed with 10X Genomics Chromium Next GEM Single Cell Multiome ATAC + Gene Expression protocol.

### Single-nucleus RNA sequencing (snRNA-seq) Library Preparation

snRNA-seq libraries were prepared using the 10x Genomics Chromium Next GEM Single Cell Multiome ATAC + Gene Expression according to the manufacturer’s protocol, with a targeted recovery of 10,000 nuclei per sample. The molarity of each library was accurately determined through qPCR utilizing the KAPA library Quantification Kit according to the manufacturer’s protocol (KAPA Biosystems/Roche) to produce cluster density appropriate for the Illumina NovaSeq X Plus instrument. Normalized libraries were sequenced on a NovaSeq X Plus Flow Cell using the151×10x24×151 sequencing recipe according to manufacturer protocol to a read depth of 500M PE reads per Gene Expression (GEX) library.

### snRNA-seq Data Processing

The reads for the 10X GEX libraries were aligned and quantitated with 10x’s CellRanger-ARC v2.0.2 against 10X’s standard refdata-cellranger-arc-GRCh38-2020-A-2.0.0 human gene reference per manufacturer’s protocol. Subsequent data analysis was performed using the Seurat R package. For the GEX analysis, low-quality cells were excluded based on three quality control metrics: the number of detected features, the proportion of mitochondrial transcripts, and total RNA counts. Specifically, we excluded cells expressing fewer than 200 or more than 7,500 genes, cells with mitochondrial transcript content exceeding 20%, and cells with fewer than 200 or more than 20,000 RNA counts. Data normalization and identification of highly variable features were performed using Seurat’s anchor-based integration strategy “SCTransform” method to integrate data across all samples. Specifically, we utilized the “PrepSCTIntegration”, “FindIntegrationAnchors”, and “IntegrateData” functions to integrate gene expression matrices from all the samples.

### Cell Type Identification

The top 3,000 highly variable genes, identified via the SCTransform workflow, were used for principal component analysis (PCA). The top 30 principal components (PCs) were selected for downstream clustering analysis based on the variance explained. Clustering was performed using Seurat’s FindNeighbors and FindClusters functions with a resolution parameter set to 0.8. The resulting clusters were visualized using Uniform Manifold Approximation and Projection (UMAP). Cell type annotation was performed based on canonical marker gene expression and gene function, guided by prior single-cell studies (*69–71*). Marker genes used for major cell types included macrophages (CD14, CD163, MS4A6A, and CSF1R), T cells (CD3D, CD3E, and CD27), meningioma cells (CLU, PTN, LEPR, and SSTR2), endothelial cells (CD34, VWF, CCL14, and PLVAP), and dendritic cells (high ITGAX and low CD14) using SCINA R package (v1.2.0). Macrophage sub-clustering was performed using the same PCA-based approach as described above. Marker genes for each resulting cluster were identified using the “PrepSCTFindMarkers” and “FindAllMarkers” functions in Seurat.

### Cell-cell Communication Network Analyses

Cell-cell communication networks analyses were perfromed using CellChat v2.0 which utilizes ligand-receptor interactions from the manually curated CellChatDB database (CellChatDB.human). Communication probability, cellular communication signaling pathways inference, and the aggregated cell-cell communication network were calculated using CellChat R package *computeCommunProb* and *computeCommunProbPathway* module. Visualization of the communication pattern and the network centrality scores were computed using *CellChat netAnalysis_computeCentrality* modeule.

### Statistical Analysis

Comparison between groups for categorical variables were performed using Fisher’s Exact test or Pearson’s chi squared test, while continuous variables were compared using Mann-Whitney U test. Correlations between continuous variables were assessed using Spearman’s rank correlation test (ρ). PFS and OS were estimated using the Kaplan-Meier method and compared using the log-rank test. All time-to-event data were calculated from the start of neoadjuvant therapy, and all statistical tests were two-sided. Statistical analyses were performed with the Statistical Package for Social Sciences, version 23.0 (IBM SPSS Statistics, Chicago, IL, USA) and GraphPad Prism version 10 (GraphPad Software, Inc., San Diego, CA, USA).

## Acknowledgements

We thank the Alvin J. Siteman Cancer Center at Washington University School of Medicine and Barnes-Jewish Hospital in St. Louis, MO, for the use of the Shared Resources including Clinical Trials Core and Tissue Procurement Core. We thank David Schwab, Konstantina Stavroulaki, Casey Hatscher, and Stephanie Myles in the Department of Radiation Oncology for clinical trial oversight and enrollment. We thank Marco Colona and Simone Brioschi for their helpful discussions and suggestions. We thank Brian Goetz from the Tissue Procurement Core. The Siteman Cancer Center is supported in part by an NCI Cancer Center Support Grant #P30 CA091842. We thank the Department of Radiation Oncology for shared resources.

## Funding

Research reported in this publication was supported in part by the institutional clinical trial grant from the department of Radiation Oncology, Washington University School of Medicine (JH). This publication was also supported in part by the Washington University Institute of Clinical and Translational Sciences which is, in part, supported by the NIH/National Center for Advancing Translational Sciences (NCATS), CTSA grant #UL1TR002345. This publication was also funded in part by the Alvin J. Siteman Cancer Center and Barnes-Jewish Hospital. The Siteman Cancer Center is supported in part by an NCI Cancer Center Support Grant #P30 CA091842. Additional support was provided by the Barnard Cancer Institute. This study was also supported by Pfizer and the healthcare business of Merck KGaA, Darmstadt, Germany (CrossRef Funder ID: 10.13039/100009945), who provided avelumab free of charge.

## Ethics approval and consent to participate

The study was approved by the Institutional Review Board and was conducted in accordance with the Declaration of Helsinki and Good Clinical Practice guidelines. All patients provided written-informed consent for participation in the study.

## Data Availability

The data generated in this study are not publicly available due to patient privacy and consent but are available upon reasonable request from the corresponding author.

## Competing interests

Tanner Johanns has served on an advisory board for Servier and Replimune. Konstantin Chernyshov and Basim Salem have BostonGene Corporation stock options. All other authors declare they have no competing interests.

## Author Contribution

Conceptualization: Jiayi Huang

Methodology: Danil Ivanov, Basim Salem, Chun-Kan Chen

Investigation: Tanner Johanns, Jian Campian, Michael Chicoine, Gregory Zipfel, Milan Chheda, Omar Butt, Albert Kim, Chun-Kan Chen

Formal Analysis: Jiayi Huang, Chun-Kan Chen, Tanner Johanns, Danil Ivanov, Konstantin Chernyshov, Basim Salem, Arina Tkachuk, Alena Frank, Daria Klimova, Dmitry Tabakov, Akdes S. Harmanci, William Chen, Akash Patel, David Raleigh,

Writing – original draft: Jiayi Huang

Writing – review and editing: Chun-Kan Chen, Tanner Johanns, Jian Campian, Michael Chicoine, Gregory Zipfel, Milan Chheda, Omar Butt, Danil Ivanov, Konstantin Chernyshov, Basim Salem, Arina Tkachuk, Alena Frank, Daria Klimova, Dmitry Tabakov, Akdes S. Harmanci, William Chen, Akash Patel, David Raleigh, Albert Kim,

Visualization: Jiayi Huang, Tanner Johanns, Chun-Kan Chen, Danil Ivanov, Konstantin Chernyshov, Basim Salem, Arina Tkachuk, Alena Frank, Daria Klimova, Dmitry Tabakov, Akdes S. Harmanci,

Supervision: Jiayi Huang

Project administration: Jiayi Huang, Danil Ivanov, Dmitry Tabakov, Chun-Kan Chen Funding acquisition: Jiayi Huang

## ClinicalTrials.gov

NCT03267836.

Presented in part as an oral presentation at the American Society of Clinical Oncology (ASCO) Annual Meeting, June 2024, Chicago, IL.

## Supplementary Materials

### Supplementary Figures

**Fig. S1.**
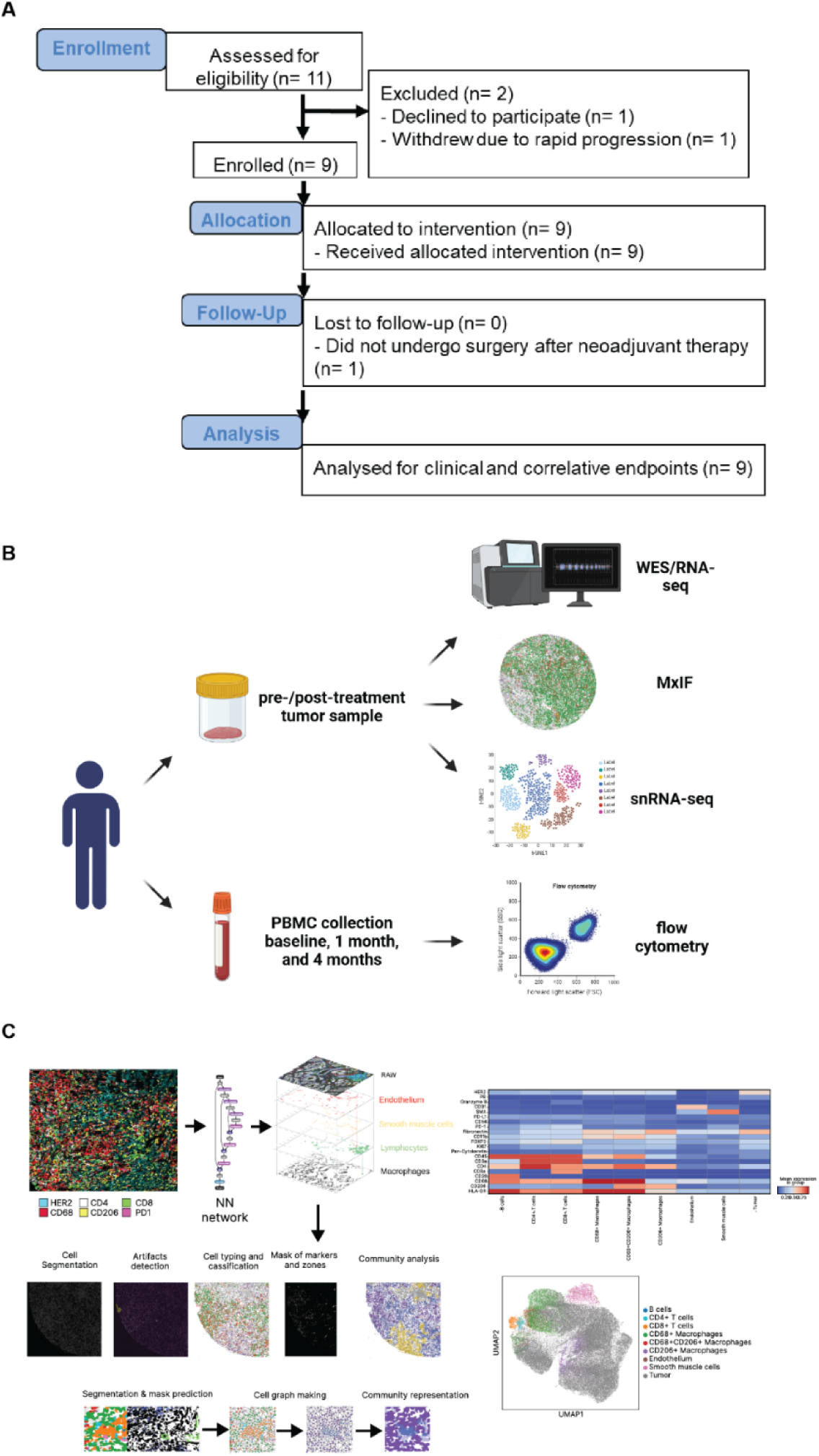
Study design. (A) (C) Consort diagram. (B) Schema of the multi-omic correlative analyses performed on the tumor samples and peripheral blood mononuclear cells (PBMCs) from this window-of-opportunity study: whole exome sequencing (WES), RNA sequencing (RNA-seq), multiplex immunofluorescence (MxIF), single-nucleus RNA sequencing (snRNA-seq), and flow cytometry. (C) Analytical workflow of multiplex immunofluorescence (MxIF).

**Fig. S2.**
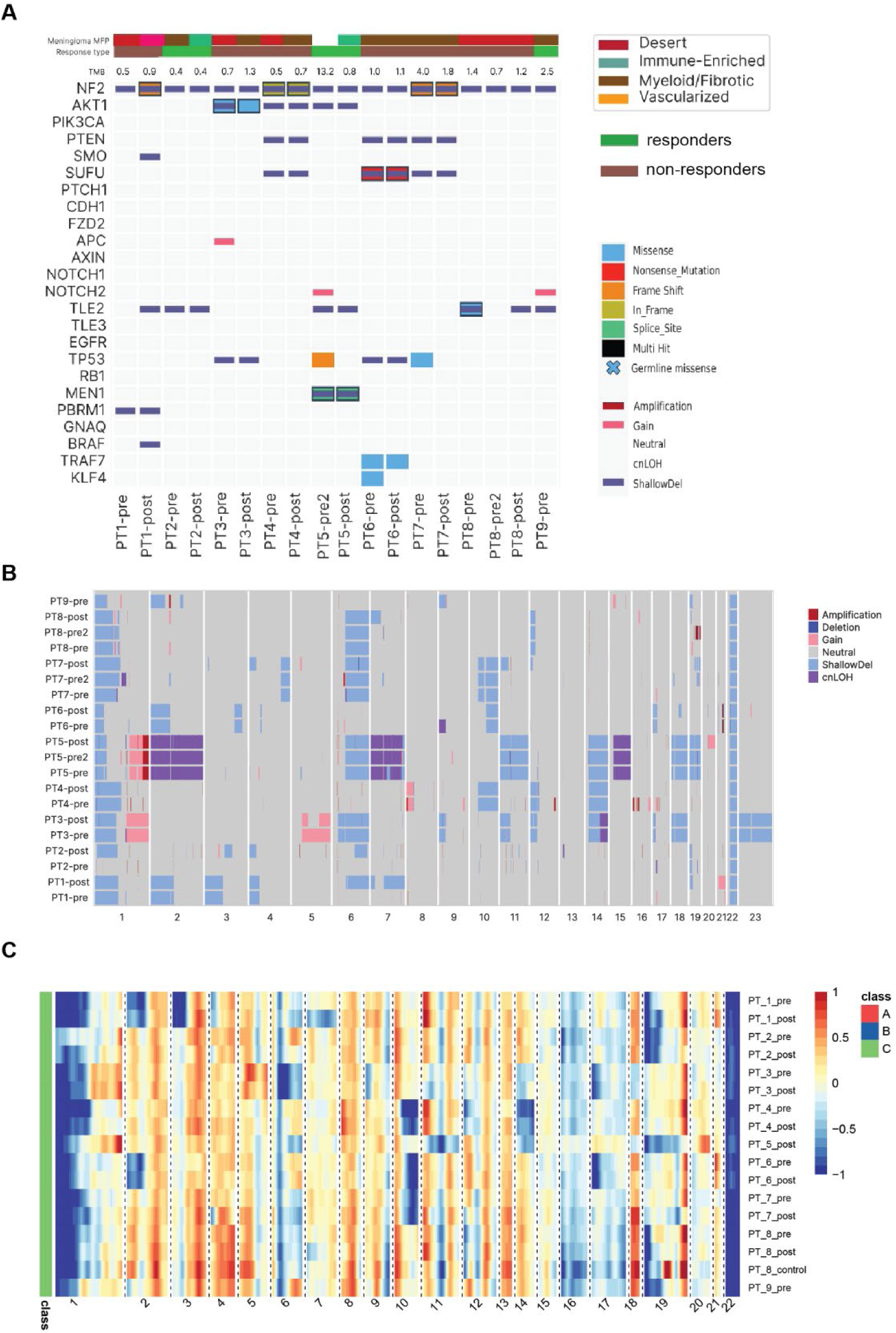
Tumor mutations and chromosome alterations. (A) Tumor mutations as assessed by whole exome sequencing (WES). Tumor mutation burden (TMB) was derived from WES to quantify mutations per megabase in the coding region of the genome. (B) Chromosomal alterations derived from WES. (C) Chromosomal alterations derived from RNA-seq.

**Fig. S3.**
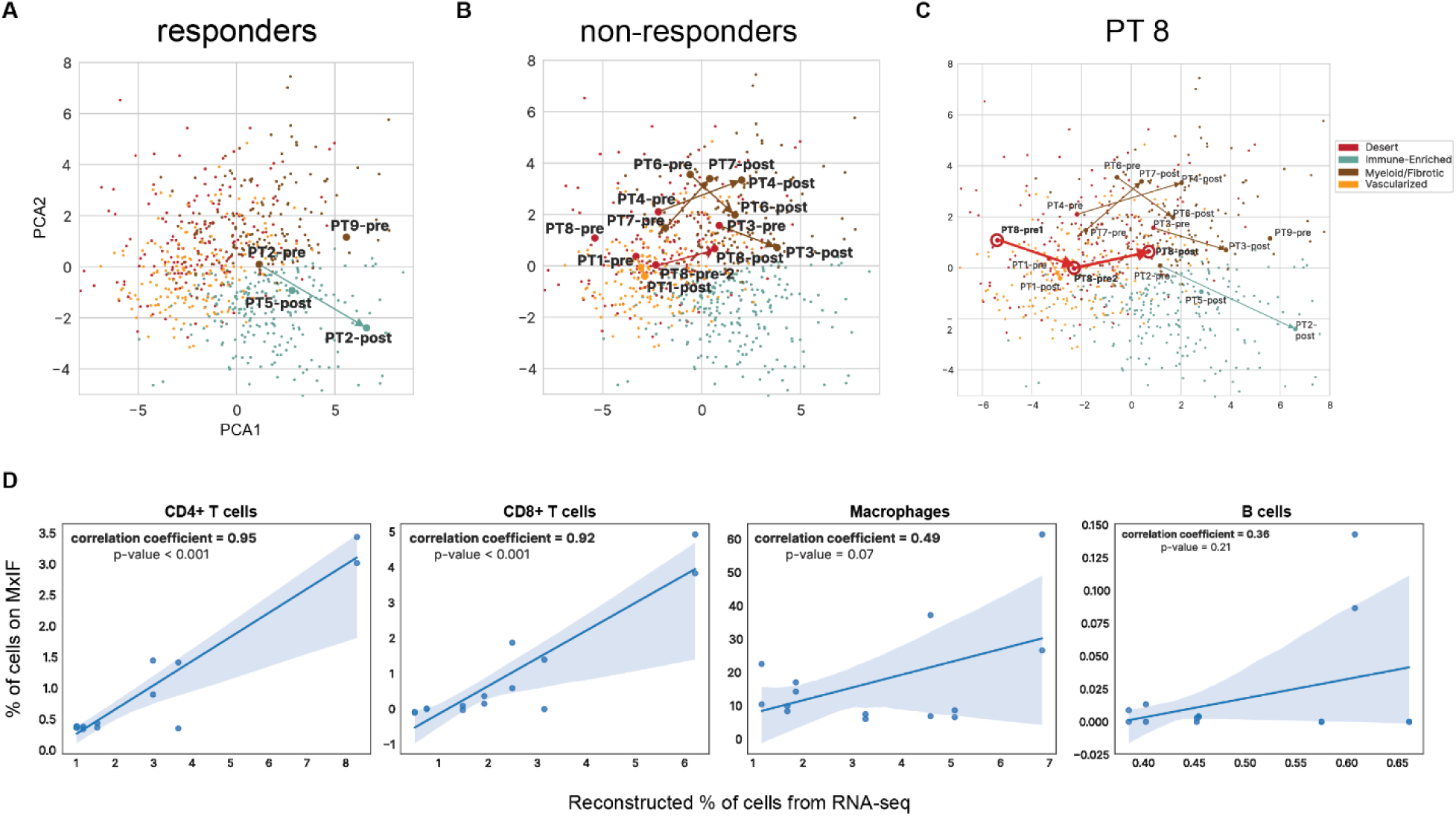
Tumor micro-environment (TME) changes. PCA plot depicting dynamic changes of MFP signatures after radio-immunotherapy in responders (A), non-responders (B), and a non-responder patient with two pre-treatment samples before and after the initial radiation therapy course (C). (D) Scatter showing correlation between percentage of specific immune cell type on multiplex immunofluorescence (MxIF) and corresponding reconstructed percentage of immune cell type derived from RNA-seq using Kassandra algorithm pre-trained on artificial transcriptomes. Two representative cores from the pre- and post-treatment tumor samples of 4 patients were analyzed with MxIF, yielding a total of 16 cores.

**Fig. S4.**
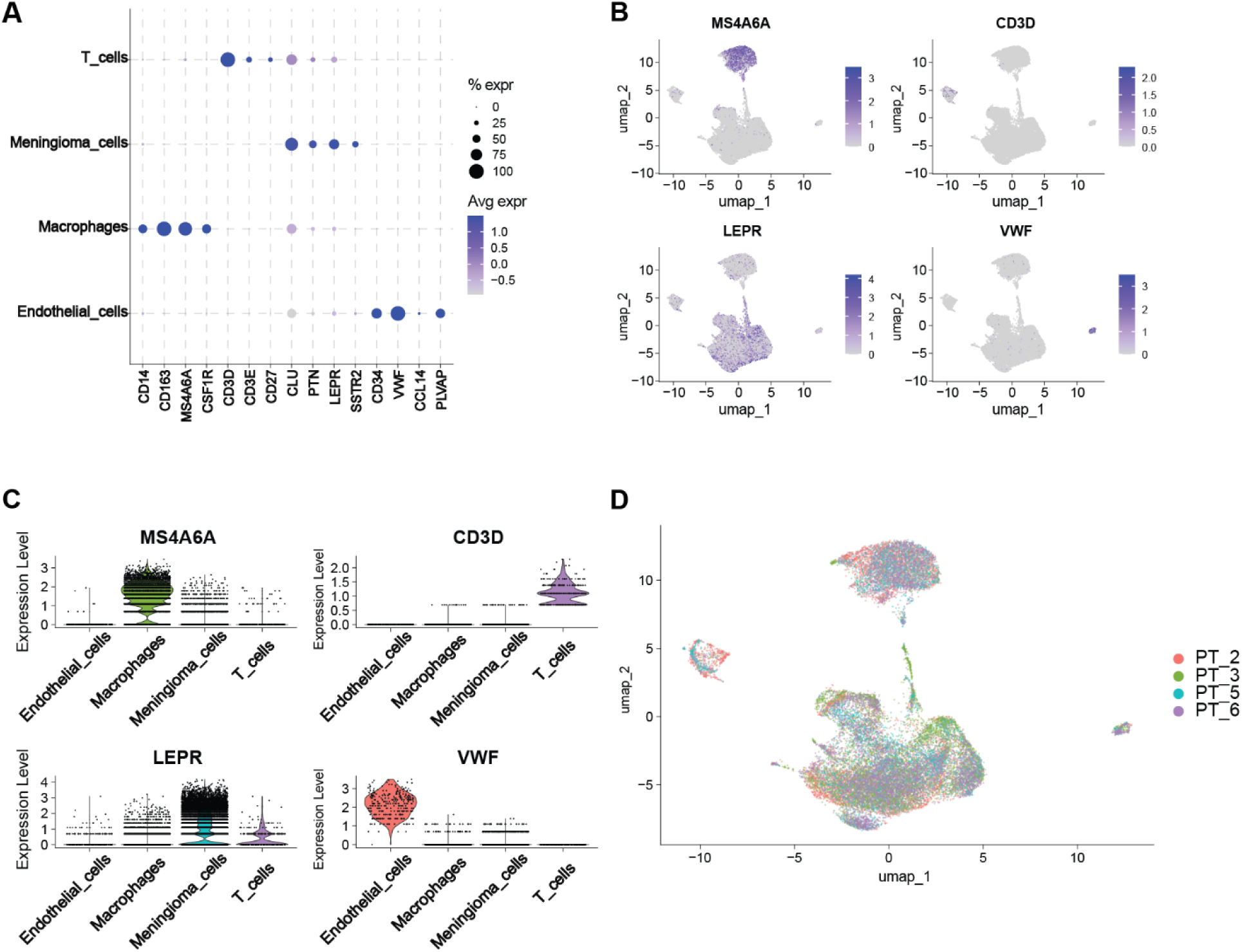
snRNA-seq analyses of the post-treatment tumor samples. **(A)** The dot plot displaying the expression of selected marker genes for each annotated cell identity. Scaled color bar = average expression, size of the point = percent expressed. **(B)** The feature plots displaying the expression of the most representative marker gene for each annotated cell identity. **(C)** violin plot showing the expression profile of a representative marker gene across all single cells, with color intensity indicating the normalized expression level of the marker gene. **(D)** UMAP plot displaying 25,983 single cells, with each point representing an individual cell and colors indicating the patient tumor sample where the cell derived from.

**Fig. S5.**
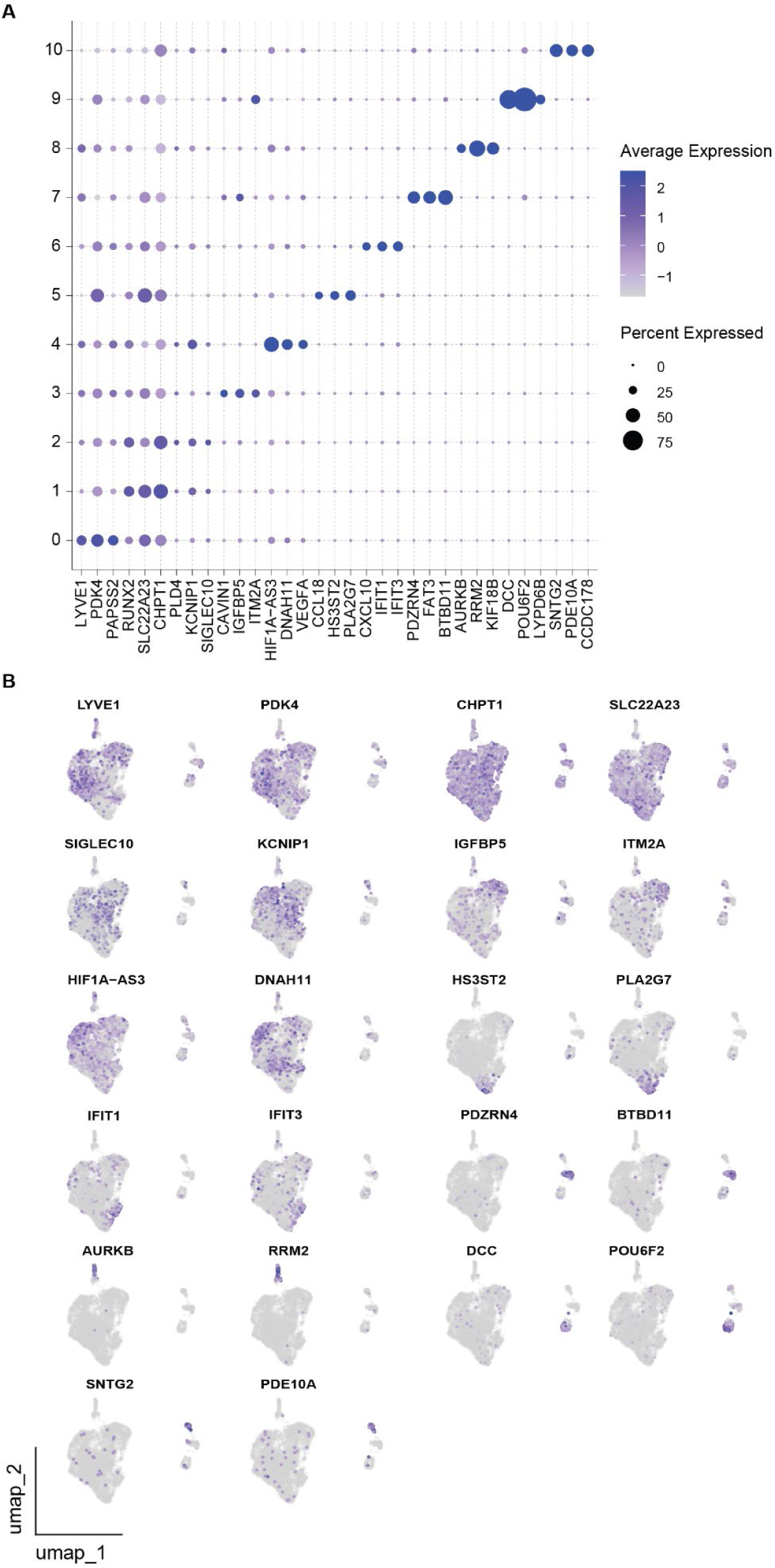
snRNA-seq analyses of the macrophages sub-clusters. **(A)** The dot plot displaying the expression of selected marker genes for each macrophage sub-cluster. Scaled color bar = average expression, size of the point = percent expressed. **(B)** The feature plots showing the expression of the marker genes for each macrophage sub-cluster.

**Fig. S6.**
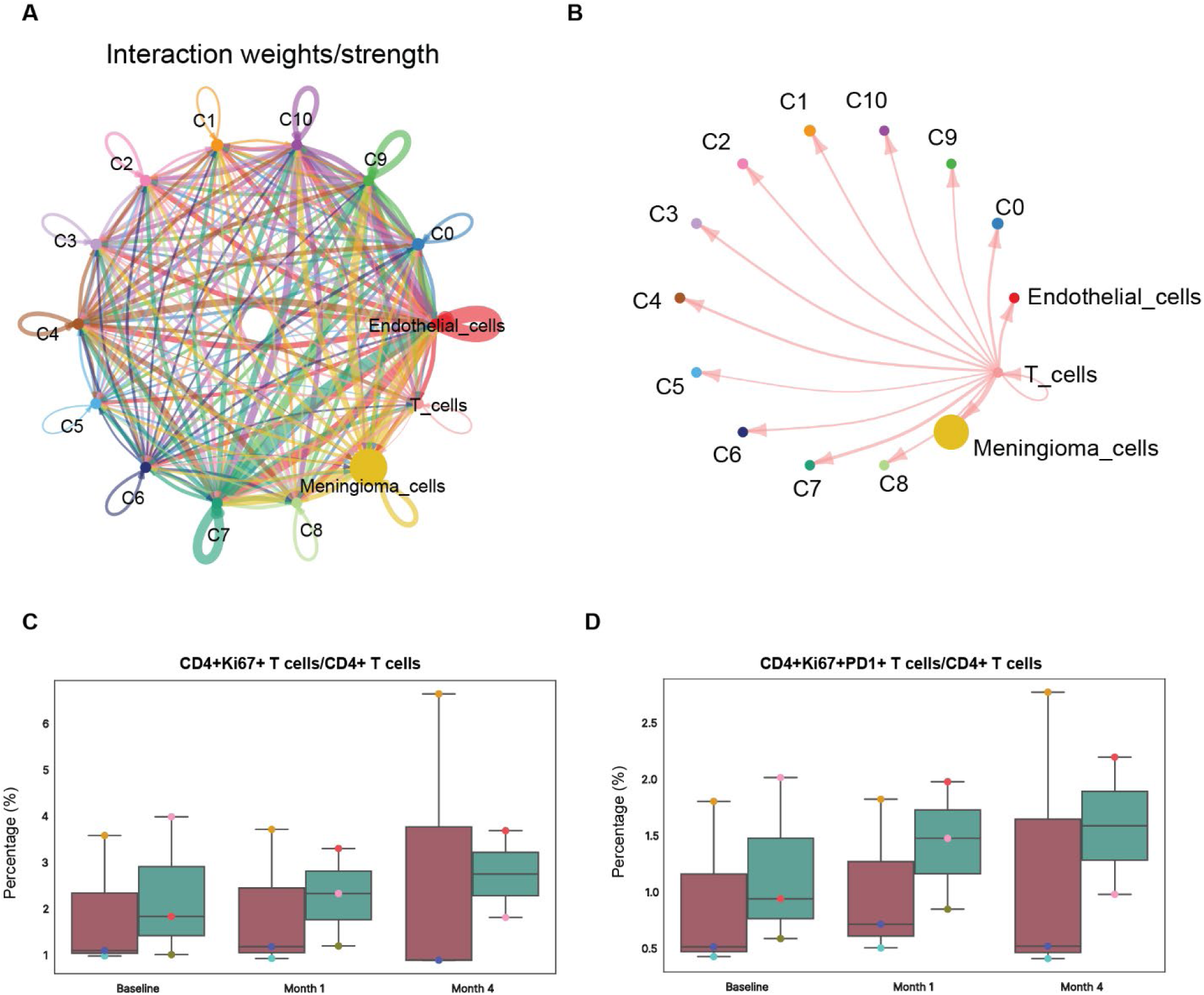
T cell interaction and blood sample analyses. **(A)** Cell-cell interaction of tumor cells with macrophage clusters, endothelial cells, and T cells. **(B)** Cell-cell interaction between T cells and the corresponding cell population. The thickness of the lines represents the weighted strength of the interaction between each corresponding cell pair. **(C-D)** Box plot comparing proportion of CD4^+^Ki67^+^ T cells or CD4^+^Ki67^+^PD1^+^ T cells among all CD4^+^ T cells between responders and non-responders before and during treatment.

### Supplementary Tables

**Supplementary Table S1.** Dosimetric Constraints.

| Dose Metric | Per Protocol | Deviation Acceptable | Deviation Unacceptable |
| --- | --- | --- | --- |
| PTV covered by 95% of prescription dose (19 CGE) | $\geq 95\%$ | $\geq 90\%$ | $< 90\%$ |
| Optic Nerves and Chiasm (0.03cc) | $\leq 20$ CGE | $\leq 24$ CGE | $> 24$ CGE |
| Brainstem (0.03 cc) | $\leq 22$ CGE | $\leq 26$ CGE | $> 26$ CGE |
| Eyes (0.03 cc) | $\leq 22$ CGE | $\leq 26$ CGE | $> 26$ CGE |
| Volume of Brain-GTV receiving 20 CGE | $\leq 20$ cc | $\leq 30$ cc | NA |
| Minimum dose to the PTV (0.03 cc) | $\geq 17$ CGE (85% of prescription dose) | $\geq 16$ CGE (80% of prescription dose) | $< 16$ CGE |
| Maximum dose to the PTV (0.03cc) | $\leq 24$ CGE (120% of prescription dose) | $\leq 26$ CGE (130% of prescription dose) | $> 26$ CGE |

**Supplementary Table S2.** Publicly available RNA-seq and microarray datasets of meningiomas.

| Dataset | Number of patients | Reference Link |
| --- | --- | --- |
| GSE74385 | 62 | https://pubmed.ncbi.nlm.nih.gov/26894859/ |
| GSE84263 | 96 | https://www.ncbi.nlm.nih.gov/pmc/articles/PMC5114141/ |
| GSE101638 | 42 | https://www.ncbi.nlm.nih.gov/pmc/articles/PMC8204688/ |
| GSE183653 | 185 | https://www.ncbi.nlm.nih.gov/pmc/articles/PMC9374001/#SD1 |
| PRJNA705586* | 64 | https://www.ncbi.nlm.nih.gov/pmc/articles/PMC9287111/ |
| EGAS00001004982 | 124 | https://www.ncbi.nlm.nih.gov/pmc/articles/PMC6131140/ |
| GSE189672* | 110 | https://www.science.org/doi/10.1126/sciadv.abm6247?url_ver=Z39.88-2003&rfr_id=ori:rid:crossref.org&rfr_dat=cr_pub%20%20pubmed |
| GSE136661† | 59 | https://www.ncbi.nlm.nih.gov/pmc/articles/PMC6815170/ |

\*All GSE189672 samples (n = 110) and a subset of PRJNA705586 (n = 28) had progression-free survival data.
†Of the 160 cases reported, 101 are the same as in GSE189672, so only 59 non-duplicate cases were used.

**Supplementary Table S3.** The antibody composition of the MxIF panel.

| Antibody | Source | Clone |
| --- | --- | --- |
| DAPI | Akoya | n/a |
| CD45 | BioLegend | 2D1 |
| CD8 | BioLegend | C8/144B |
| CD4 | Akoya | EPR6855 |
| FoxP3 | Akoya | 236A/E7 |
| CD3 | Akoya | EP449E |
| CD20 | Akoya | L26 |
| CD68 | Akoya | KP1 |
| CD11b | CST | D6X1N |
| HLA-DR | Akoya | EPR3692 |
| CD11c | Akoya | 118/A5 |
| GZB | Akoya | D6E9W |
| PD1 | Akoya | D4W2J |
| CD206 | Bethyl/Fortis | BLR109H |
| CD56 | Akoya | CAL53 |
| Ki67 | Akoya | B56 |
| PD L1 | Akoya | 73-10 |
| PanCK | Akoya | AE-1/AE-3 |
| NaK ATPase | abcam | EP1845Y |
| SMA | Akoya | 1A4 |
| Fibronectin | abcam | F1 |
| Vimentin | Akoya | 091D3 |
| CD31 | Akoya | EP3095 |
| ER | abcam | SP1 |
| PR | abcam | SP2 |
| Her2 | Bethyl | BLR241L |

**Supplementary Table S4.** Major cell populations from MxIF.

| Cell type name | Major population category | Phenotype |
| --- | --- | --- |
| B cells | B cells | CD45+ CD20+ CD3- |
| CD4+ T cells | CD4+ T cells | CD45+ CD3+ CD4+ |
| CD8+ T Cells | CD8+ T Cells | CD45+ CD3+ CD8+ |
| CD68+ Macrophages | CD68+ Macrophages | CD68+ CD206- |
| CD206+ Macrophages | CD206+ Macrophages | CD68- CD206+ |
| CD68+CD206+ Macrophages | CD68+CD206+ Macrophages | CD68+ CD206+ |
| Endothelium | Endothelium | CD31+ SMA+/- |
| Smooth muscle cells | Smooth muscle cells | SMA+ CD31- |
| Necrosis | Necrosis | In this case unique combination of markers were expressed in necrotic tissue:<br>Fibronectin+CD11b+<br>Other_markers- |
| Tumor | Tumor cells | Mix of contradictory markers or no expression at all except DAPI.<br>Tumor cells were assessed by pathology analyst to validate the population definition |
| T helpers | CD4+ T cells | CD45+ CD3+ CD4+ FoxP3- |
| T regs | CD4+ T cells | CD45+ CD3+ CD4+ FoxP3+ |
| PD-1+ T helpers | CD4+ T cells | CD45+ CD3+ CD4+ FoxP3- PD-1+ |
| PD-1+ T regs | CD4+ T cells | CD45+ CD3+ CD4+ FoxP3+ PD-1+ |
| GranzymeB+ CD8+ T cells | CD8+ T Cells | CD45+ CD3+ CD8+ GranzymeB+ |
| Ki67+ CD8+ T cells | CD8+ T Cells | CD45+ CD3+ CD8+ Ki67+ |
| PD-1+ CD8+ T cells | CD8+ T Cells | CD45+ CD3+ CD8+ PD-1+ |
| PD-L1+ CD8+ T cells | CD8+ T Cells | CD45+ CD3+ CD8+ PD-L1+ |
| HLA-DR+ CD68+ Macrophages | CD68+ Macrophages | CD68+ CD206- HLA-DR+ |
| Ki67+ CD68+ Macrophages | CD68+ Macrophages | CD68+ CD206- Ki67+ |
| PD-1+ CD68+ Macrophages | CD68+ Macrophages | CD68+ CD206- PD-1+ |
| PD-L1+ CD68+ Macrophages | CD68+ Macrophages | CD68+ CD206- PD-L1+ |
| HLA-DR+ CD206+ Macrophages | CD206+ Macrophages | CD68- CD206+ HLA-DR+ |
| Ki67+ CD206+ Macrophages | CD206+ Macrophages | CD68- CD206+ Ki67+ |
| PD-L1+ CD206+ Macrophages | CD206+ Macrophages | CD68- CD206+ PD-L1+ |
| HLA-DR+ CD68+CD206+ Macrophages | CD68+CD206+ Macrophages | CD68+ CD206+ HLA-DR+ |
| PD-1+ CD68+CD206+ Macrophages | CD68+CD206+ Macrophages | CD68+ CD206+ PD-1+ |
| PD-L1+ CD68+CD206+ Macrophages | CD68+CD206+ Macrophages | CD68+ CD206+ PD-L1+ |
| Ki67+ CD68+CD206+ Macrophages | CD68+CD206+ Macrophages | CD68+ CD206+ Ki67+ |
| Tumor HER2+ | Tumor cells | HER2+ |
| PR+ Tumor | Tumor cells | PR+ |
| Tumor HER2+ PR+ | Tumor cells | HER2+PR+ |
| Fibronectin+ Tumor cells | Tumor cells | Fibronectin+ |
| Fibronectin+ HER2+ Tumor cells | Tumor cells | Fibronectin+HER2+ |
| PR+ Fibronectin+ Tumor cells | Tumor cells | Fibronectin+PR+ |
| Ki67+ Tumor | Tumor cells | Ki67+ |

**Supplementary Table S5.** Defined cellular communities with description and compositions of cell populations within the community.

| Community cluster name | Community description | Cell population composition |
| --- | --- | --- |
| Predominance of tumor | Cellular community is characterized by a high concentration of tumor cells, with minimal presence of other cell types. The tumor cells dominate the cellular landscape. | 95% Tumor cells<br>5% Other populations (immune and stromal cells) |
| Tumor region with the predominance of CD206+ macrophages | Cellular community is characterized by a high concentration of CD206+ macrophages in the tumor zone, which are typically associated with tumor-promoting activities. These macrophages play a role in creating an immunosuppressive environment, suggesting a dynamic interaction between immune cells and tumor cells that promotes tumor survival and progression. | 68% Tumor cells<br>31% CD206+ Macrophages<br>1 % Other populations (immune and stromal cells) |
| Tumor region with a presence of CD68+ macrophages | Cellular community is characterized by a tumor region with a presence of CD68+ macrophages. The predominance of CD68+ macrophages and the presence of a small portion of CD68+CD206+ macrophages may suggest a complex interplay between pro-inflammatory | 48% Tumor cells<br>40% CD68+ Macrophages<br>7 % CD68+CD206+ Macrophages<br>5 % Other populations (immune and stromal cells) |
|  | and immunosuppressive processes within the tumor region. |  |
| Tumor region with the predominance of CD68+ macrophages | Cellular community characterized by a tumor region with a significant presence of CD68+ macrophages, indicating an active immune response. This community not only includes various tumor cells but also exhibits a dense infiltration of CD68+ macrophages, which are key players in the immune system. | 55% CD68<sup>+</sup> Macrophages<br>40% Tumor cells<br>5 % Other populations (immune and stromal cells) |
| Tumor-infiltrating lymphocytes (TILs) and myeloid cells within the tumor region | Cellular community is characterized by the presence of tumor-infiltrating lymphocytes (TILs) and myeloid cells within the tumor microenvironment. This community reflects the complex interplay between the immune system and the tumor, influencing tumor growth, response to therapy, and overall patient prognosis. | 40% Tumor cells<br>15% CD8<sup>+</sup> cells<br>12% CD68<sup>+</sup> Macrophages<br>11% CD68<sup>+</sup>CD206<sup>+</sup> Macrophages<br>11% CD4<sup>+</sup> cells<br>7% CD206<sup>+</sup> Macrophages<br>4% Stromal cells (Endothelium and Smooth muscle cells) |
| Blood vessels within the tumor region | Cellular community is characterized by the presence of blood vessels interspersed throughout the tumor region. | 75% Smooth muscle cells<br>20% Tumor cells<br>3% Immune cells<br>2% Endothelium |

**Supplementary Table S6.**
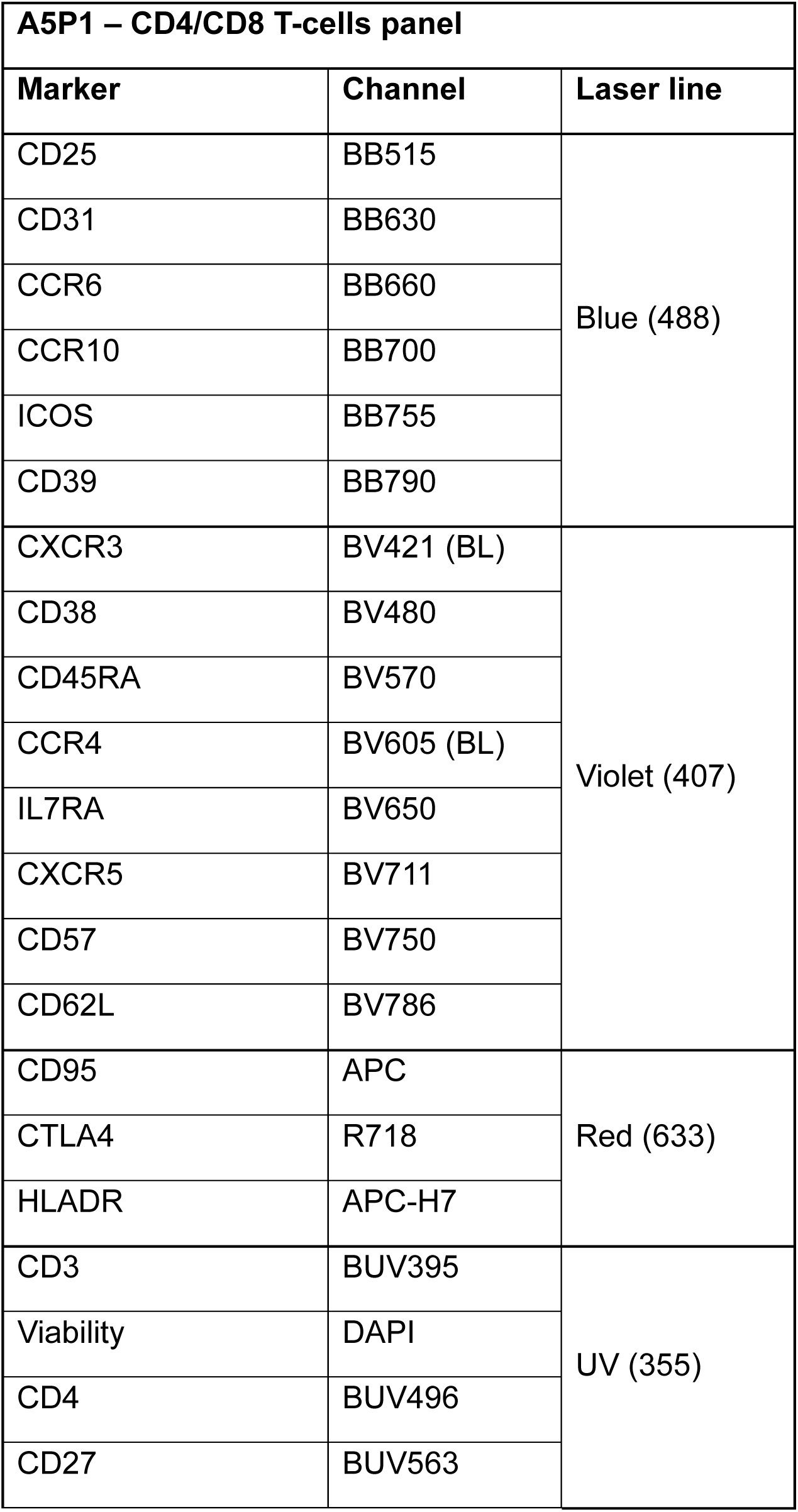
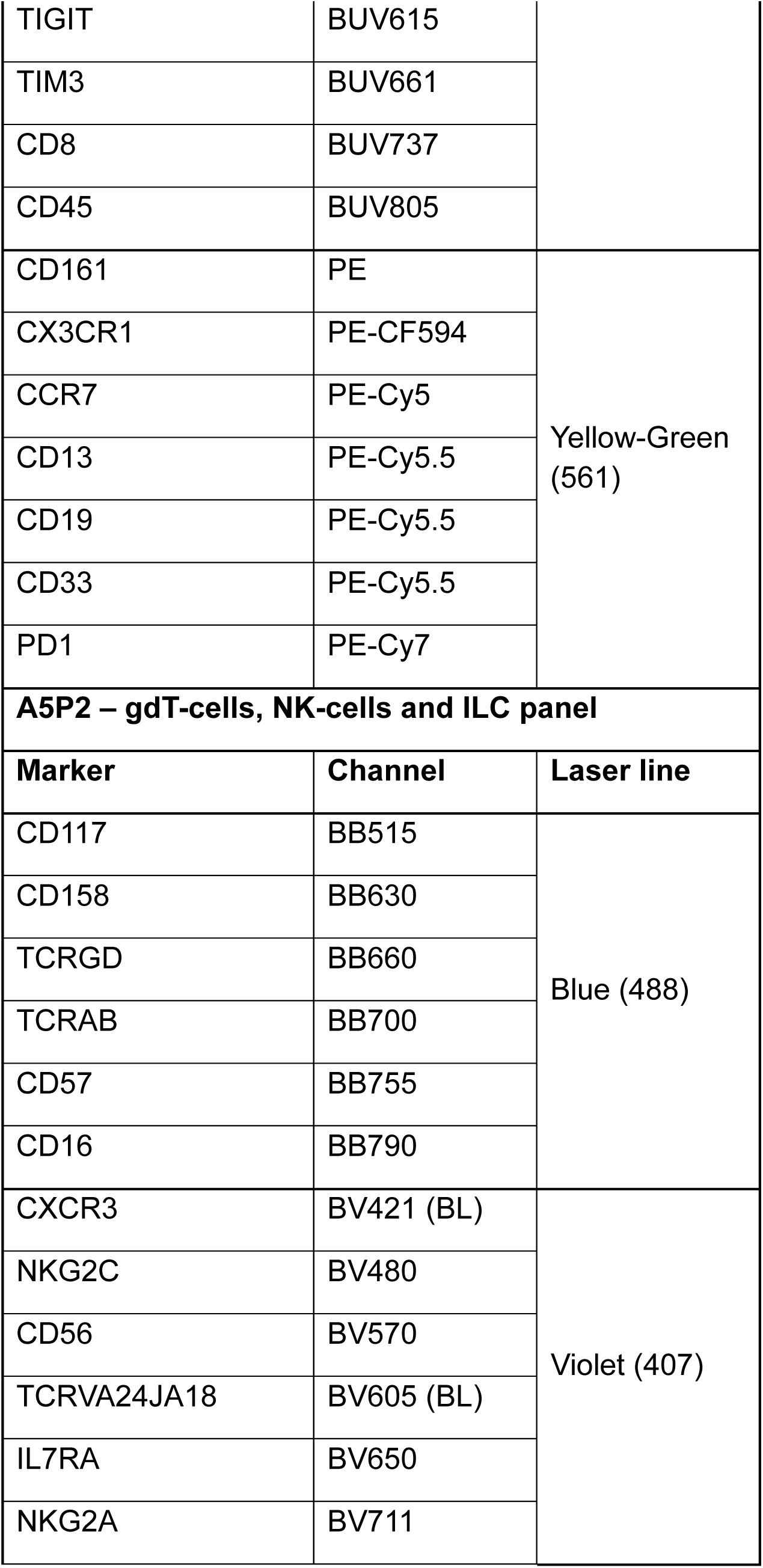
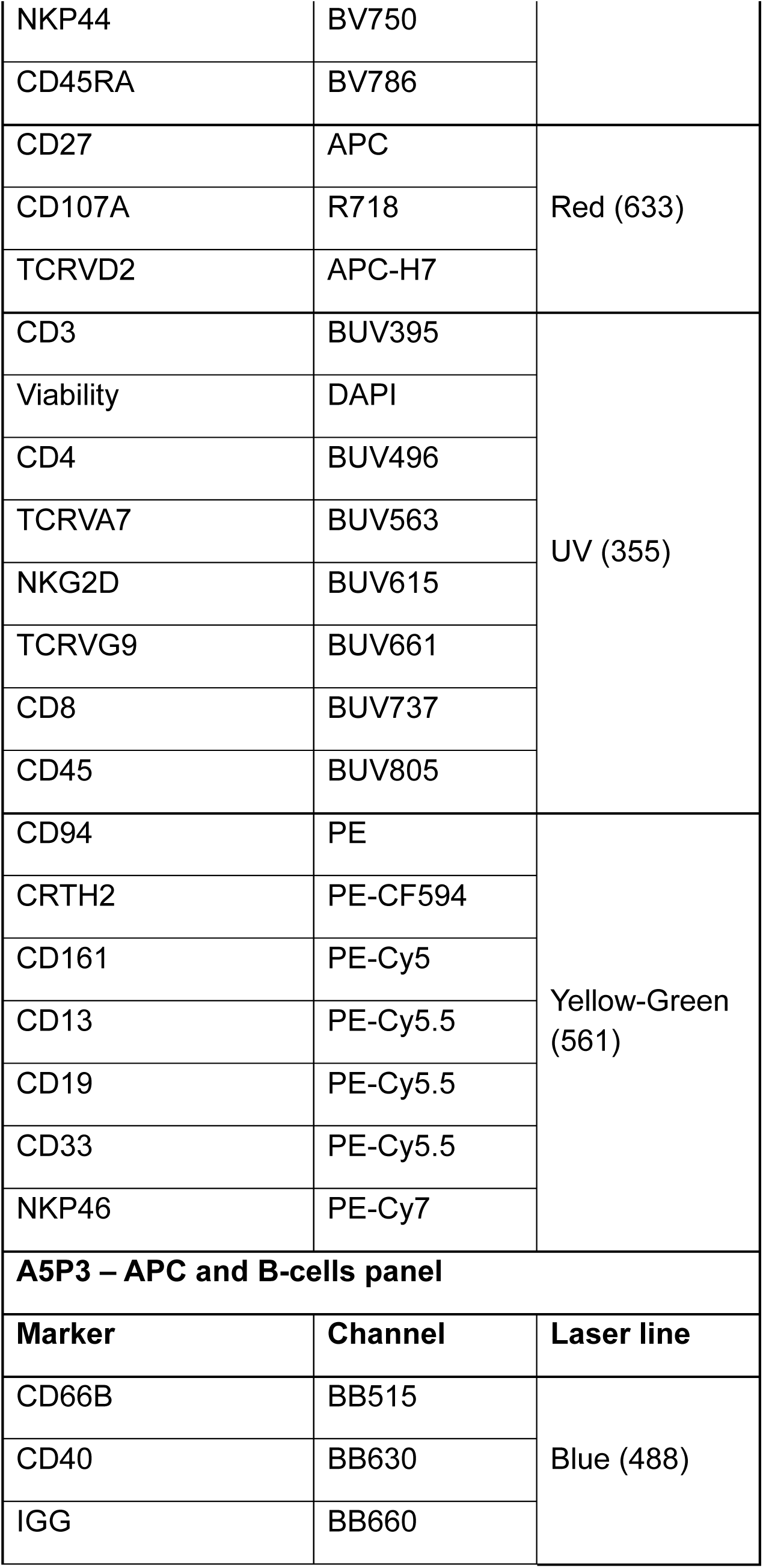
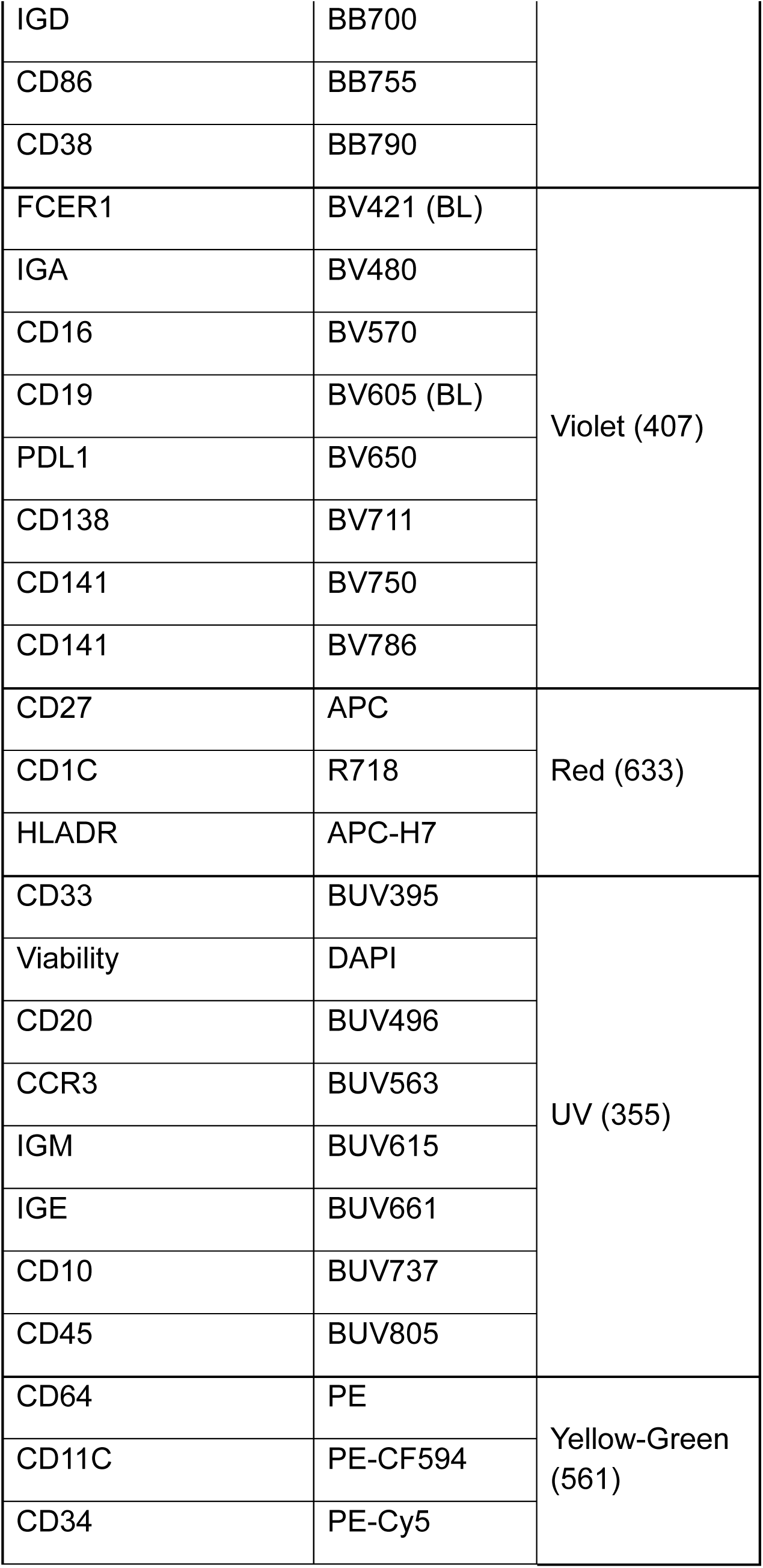
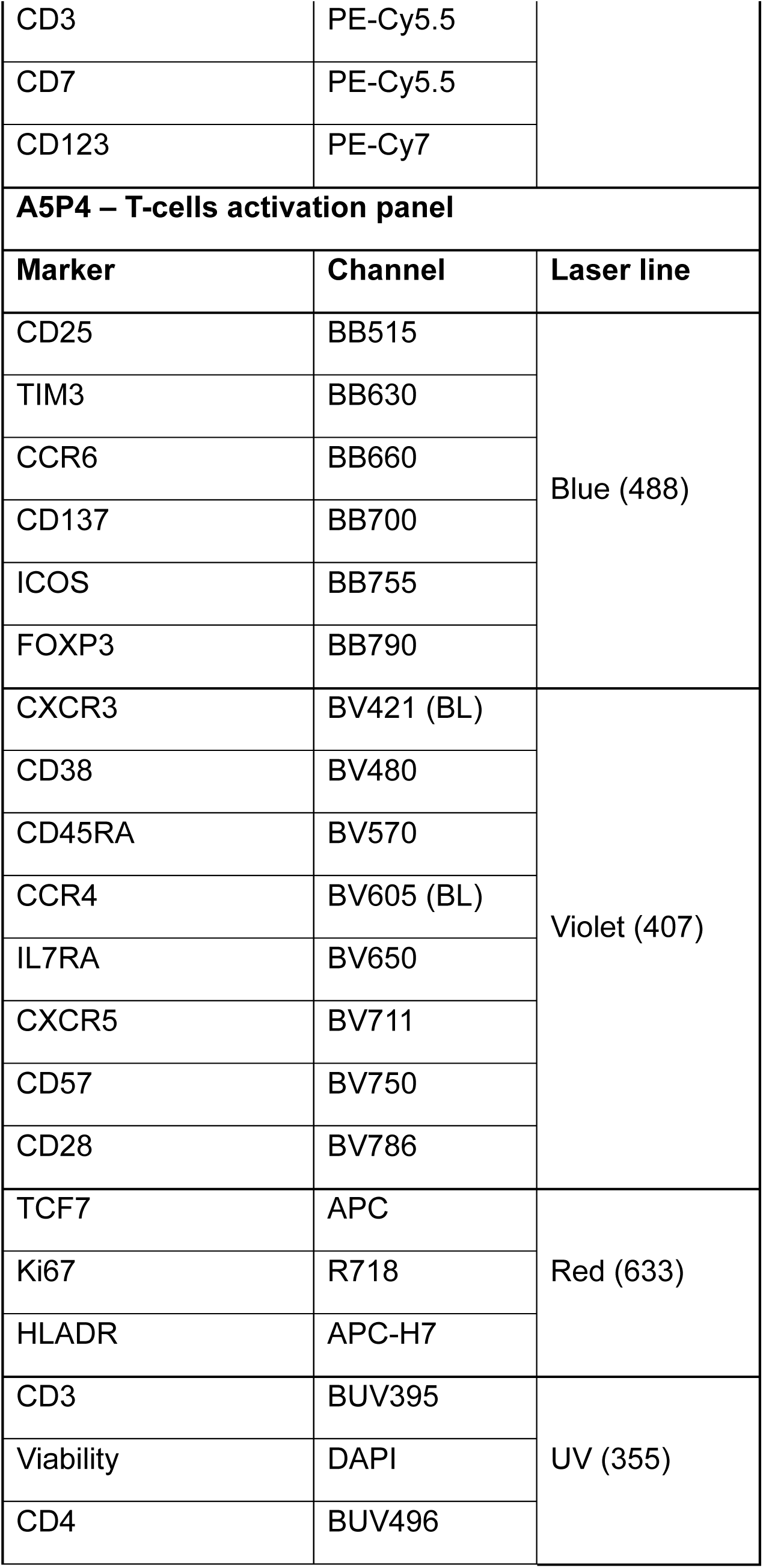
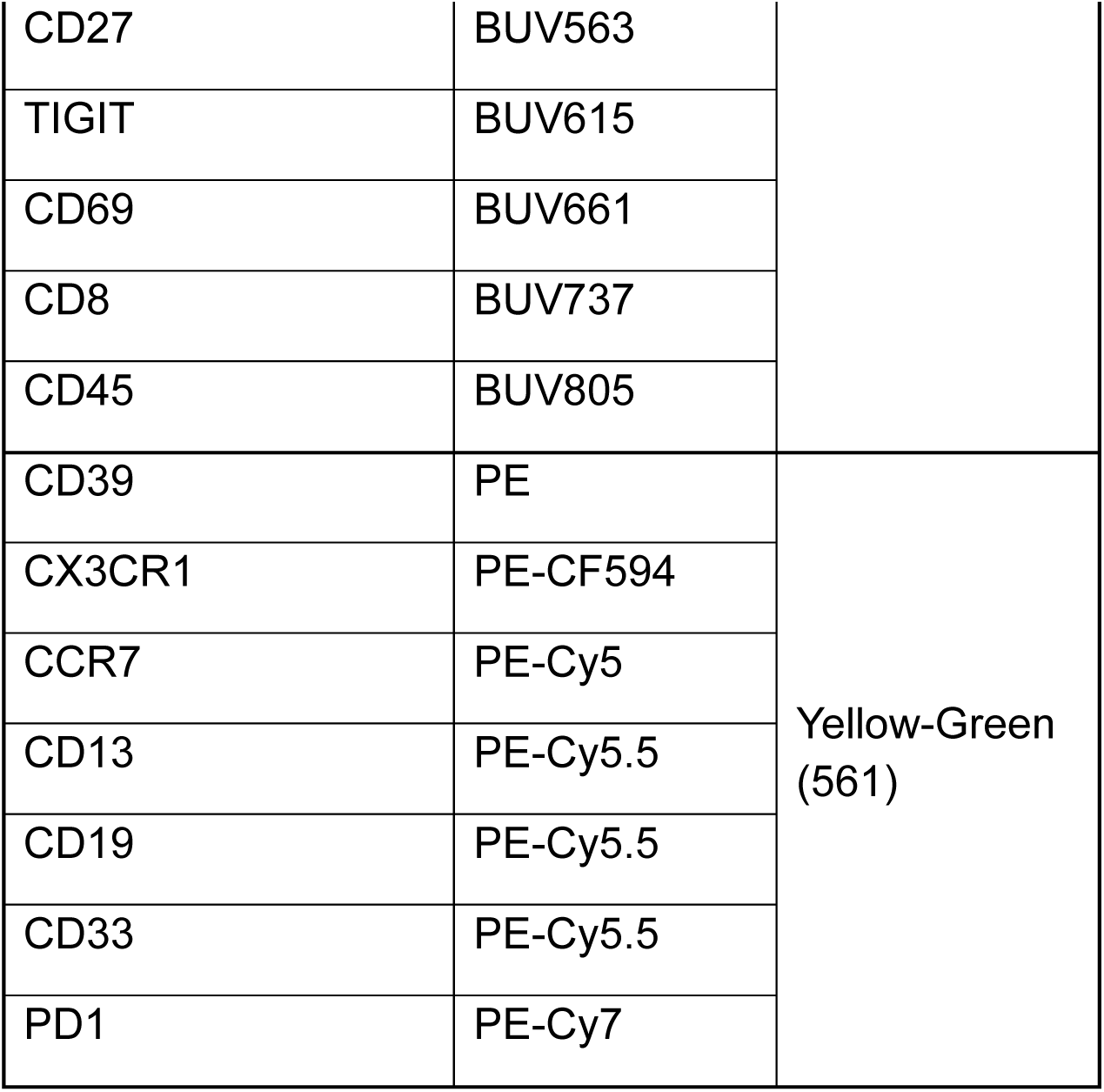
Antibody panel for flow cytometry of PBMCs.

## Notes

### Clinical Trial

NCT03267836

### Author Declarations

The study was approved by the Institutional Review Board at Washington University in St. Louis and was conducted in accordance with the Declaration of Helsinki and Good Clinical Practice guidelines. All patients provided written-informed consent for participation in the study.

